# DEPRESSION ASSOCIATED WITH INCIDENT TYPE I MI AMONG PEOPLE WITH HIV

**DOI:** 10.64898/2025.12.19.25342485

**Authors:** Emily P. Hyle, Elizabeth Humes, Acadia Thielking, Shibani S. Mukerji, Sally B. Coburn, Heidi M. Crane, Anjali Srinivasan, Kelly Gebo, Maile Karris, Natalie Pineda, Raynell Lang, David Sosa, Vincent C. Marconi, Richard D. Moore, Peter F. Rebeiro, Michael A. Horberg, Catherine R. Lesko, Sonia Napravnik, Michael J. Silverberg, Leah H. Rubin, Virginia A. Triant, Keri N. Althoff

## Abstract

**Background:** Depression and anxiety have been associated with increased risk of myocardial infarction (MI) in the general population and among people with HIV (PWH) but with limited attention to MI type. We examined the association between depression and/or anxiety and incident Type 1 (T1MI) or Type 2 (T2MI) MI among PWH.

**Methods:** We examined data from seven NA-ACCORD clinical cohorts (1997-2019) with adjudicated first MI; outcomes included T1MI (plaque rupture or cardiac intervention) or T2MI (demand ischemia). We defined depression or anxiety as a time-varying ICD-9/10-coded diagnosis prior to incident MI. We censored participants at death, disengagement from care, or first MI (if not the outcome of interest). We used Cox proportional hazard models to estimate the association between depression or anxiety and MI by type, adjusting for demographics and risk factors for MI.

**Results:** Of the 32,358 study participants, 13,751 (42.5%) had a depression diagnosis, 9,132 (28.2%) had an anxiety diagnosis, and 15,970 (47.3%) never had diagnosed depression or anxiety. After adjusting for MI risk factors, depression was associated with T1MI (aHR, 1.22 [95% CI, 1.00-1.48]), and anxiety had a protective association (albeit not statistically significant) with T1MI (aHR, 0.86 [95% CI, 0.70-1.07]). Depression had a null association (aHR, 1.05 [95% CI, 0.83-1.33] with T2MI, and anxiety was non-significantly associated with T2MI (aHR, 1.16 [95% CI, 0.89-1.51]).

**Conclusions:** Diagnosed depression was associated with T1MI but not T2MI, whereas anxiety was not statistically significantly associated with either MI type. Mental health diagnosis and treatment may play an important role in cardiovascular health among PWH.

## INTRODUCTION

In the United States, the prevalence of mood and anxiety disorders is higher among people with HIV (PWH) compared to the general population; approximately 1 in 2 PWH are diagnosed with at least one mental health disorder, 39% with depressive disorders and 28% with anxiety disorders [1–5]. Depression and anxiety are known risk factors for cardiovascular disease incidence and mortality in the general population [6–13]. Among PWH, depression has been associated with a 30% increase in acute myocardial infarction (MI) incidence and an 18% increase in stroke risk [14,15]. PWH who are diagnosed with depression and/or anxiety also often have a higher prevalence of traditional risk factors for cardiovascular disease, such as tobacco use and hypertension [16].

While the association of depression and anxiety with MI incidence has been described both among the general population and PWH, variability by type 1 MI (T1MI) or type 2 MI (T2MI) is not yet understood. T1MI, caused by plaque rupture or requiring a cardiac intervention, is associated with older age and traditional risk factors, whereas T2MI, driven by demand ischemia, can be associated with sepsis or cocaine use [17]. In contrast to T2MI, T1MI has been associated with higher CD4 count and antiretroviral therapy (ART) in PWH; however, increased mortality has been noted with T2MI compared with T1MI [18,19].

Given differences in causes, demographics, and prognosis of T1MI and T2MI, it is important to evaluate the association of depression or anxiety with each MI type among PWH to inform tailored, data-driven guidelines that integrate heart and brain health for PWH. In this analysis, we examined the relationship between diagnosis with depression and/or anxiety and T1MI or T2MI incidence among PWH in the United States.

## METHODS

### Study Population

The North American AIDS Cohort Collaboration on Research and Design (NA-ACCORD) consolidates data across 27 clinical and interval cohorts in the US and Canada [20]. The study population for this analysis includes PWH ≥18 years of age at enrollment in 7 of the NA-ACCORD clinical cohorts with observation of MI events between 1997 and 2019 (Supplementary Figure 1), which were adjudicated as previously described [3,20]. Participants without a CD4 count or HIV-1 plasma RNA viral load (VL), who never initiated ART, or who were diagnosed with bipolar disease [21], were excluded from the analysis. Study exit occurred with the first of: T1MI, T2MI, death, the conclusion of the MI observation window, or disengagement from care as defined by 2 years without an encounter or CD4 or VL testing [21].

### Outcome of Interest

The primary outcome of interest was adjudicated, incident T1MI or T2MI. A standardized protocol was used to ascertain MIs within the NA-ACCORD that was adapted from the Multi-Ethnic Study of Atherosclerosis (MESA) study and the universal MI definition, as described previously [20,22]. Participant clinical records were first screened for an inpatient or outpatient MI diagnosis (as per ICD-9/10 codes, including 410 codes, 411.0, 412, and 429.7) or serum cardiac enzyme levels (i.e., troponin-I, troponin-T, or creatine kinase MB) greater than normal. T1MI includes events caused by atherosclerotic plaque rupture, erosion with intraluminal thrombus, or requiring cardiac intervention, whereas T2MI includes events caused by an ischemic supply and demand imbalance. For all individuals identified in screening, medical records were deidentified, uploaded to a secure website, and adjudicated by two physicians with discrepancies resolved by a third reviewer.

### Exposure of Interest

The primary exposure of interest was whether participants had ever been diagnosed with depression or anxiety. Many HIV clinical sites that contribute data to NA-ACCORD conduct screening and provide treatment for mental health disorders. Diagnoses of depression or anxiety were identified using ICD-9 and ICD-10 codes in health records, as described previously [3].

### Covariates of Interest

#### Time-fixed variables

We evaluated demographic factors, including age at baseline, sex at birth, race/ethnicity, and self-reported HIV transmission risk group (i.e., heterosexual contact, men who have sex with men (MSM), and people who inject drugs [PWID]). We accounted for ever/never use of substances, including tobacco (based on diagnosis and/or substance survey data) and at-risk alcohol, ever cocaine, and ever cannabis use (based on diagnosis data). HIV-related CVD risk factors included CD4 count and VL at ART initiation, clinical AIDS diagnosis, and HCV infection (based on lab data). Supplementary Table 2 summarizes the timing of variable measurement.

#### Time-varying variables

Time-varying variables are summarized in Supplementary Table 3. We evaluated traditional CVD risk factors, including body mass index (BMI); treated hypertension (i.e., hypertension diagnosis and anti-hypertensive medication prescription); elevated total cholesterol (i.e., total cholesterol measurement ≥240 mg/dL); statin use (i.e., having an HMG-CoA reductase inhibitor prescription); chronic kidney disease (i.e., estimated glomerular filtration rate [eGFR] <60 mL/min/1.73m^2^ for at least three months based on the Chronic Kidney Disease Epidemiology Collaboration [CKD-EPI] equations); and diabetes mellitus (i.e., diabetes diagnosis and prescription of diabetes-related, or diabetes-specific medications, or glycosylated hemoglobin [HbA1c] level >6.5%). HIV-related risk factors included ART regimen (protease inhibitor (PI) vs non-PI; abacavir vs non-abacavir), CD4, and VL.

### Statistical Analysis

Multiple imputation was used to account for missing data, including tobacco use (14% missing), CD4 at ART initiation (25% missing), VL at ART initiation (27% missing), and Hepatitis C status (6% missing); the MICE package in R was used to impute 40 datasets, with logistic regression for smoking and HCV and predictive mean matching for CD4 and VL. Variables used for the imputation model included age, sex at birth, race/ethnicity, HIV transmission risk group, ever tobacco use, at risk alcohol use, ever cocaine use, ever cannabis use, hypertension, total cholesterol or statin use, chronic kidney disease, diabetes, history of AIDS diagnosis, ever HCV infection, depression, anxiety, as well as T1MI or T2MI event and cohort within NA-ACCORD.

Kaplan-Meier plots estimated the cumulative incidence of T1MI and T2MI, by those with (vs. without) a depression diagnosis and those with (vs. without) an anxiety diagnosis; a log rank test was used to estimate the p-value for a difference in curves. We used a Cox proportional hazards model adjusted for demographics, traditional CVD-risk factors, and HIV-related risk factors to estimate the relative hazards of each outcome by exposure status (the presence of depression, anxiety, both, or neither).

We tested the hypothesis that the relationship of depression and T1MI was different in PWH with and without anxiety using a nested model approach (with and without the interaction term for depression and anxiety) to determine if the model with the interaction fit better than the model without, using the multivariate Wald test. We also created a 4-category variable for depression and anxiety and estimated the relationships of depression only, anxiety only, or both depression and anxiety (vs. neither depression or anxiety) with T1MI. We then repeated this approach for the T2MI outcome. We also assessed two additional adjusted models: one that omitted depression, and one that omitted anxiety. To assess if associations between depression, anxiety, and type of MI were different in males or females, we performed sex-stratified analyses.

## RESULTS

Of 41,071 PWH aged 18 years or older at enrollment in NA-ACCORD, 4,743 never initiated ART, 1 never had a CD4 or VL, and 3,969 had a bipolar disorder diagnosis. The remaining 32,358 participants contributed 176,310.3 cumulative person-years. Among participants, 13,751 (42.5%) had a depression diagnosis, and 9,132 (28.2%) had an anxiety diagnosis (Table 1); 6,495 (20.1%) had both a depression and anxiety diagnosis (Supplementary Table 1). Most participants were male (83.5%), non-Hispanic White (45.0%), and identified as MSM (57.7%). Median age at baseline was 42 years (IQR, 34-49).

**Table 1.**
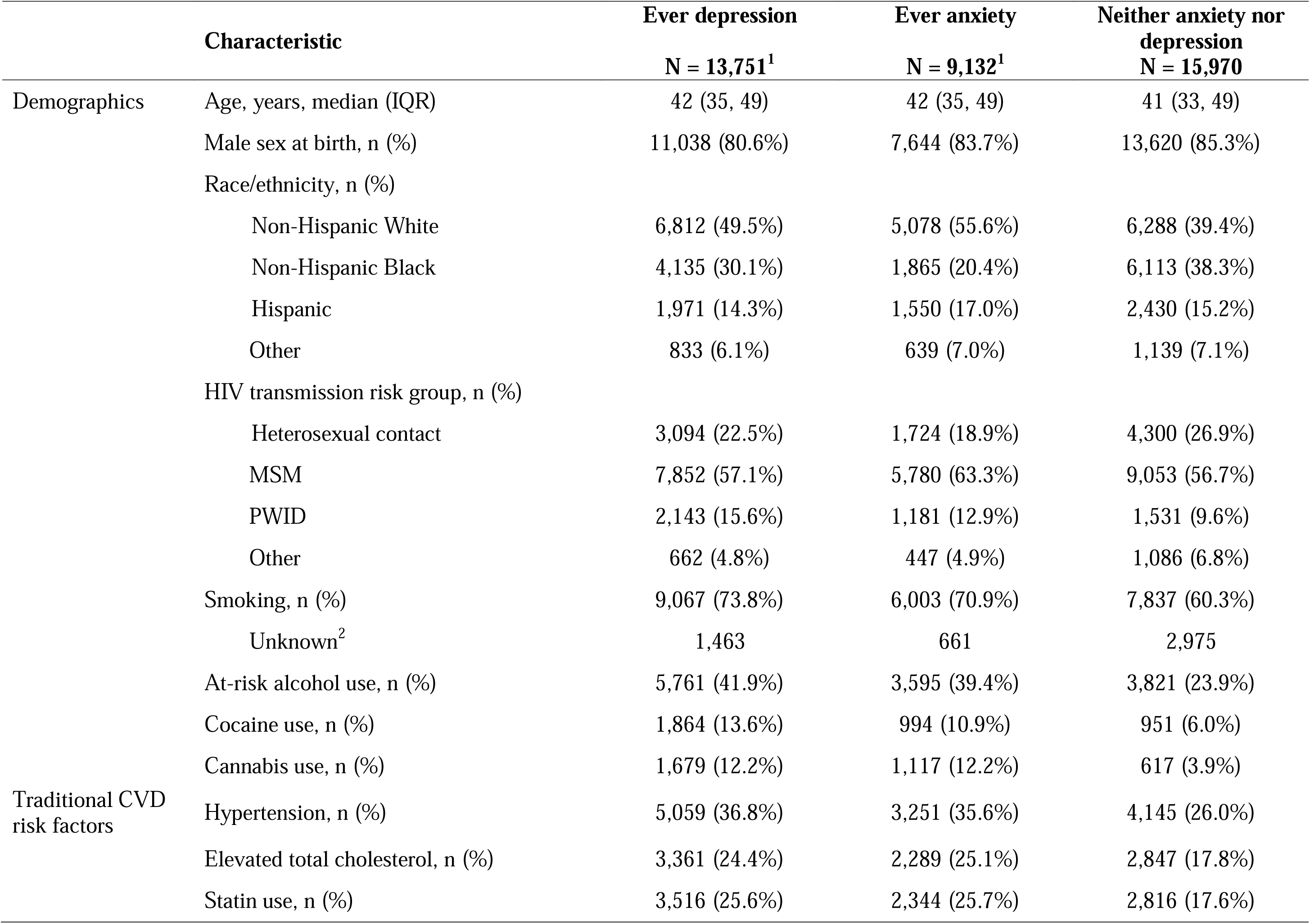
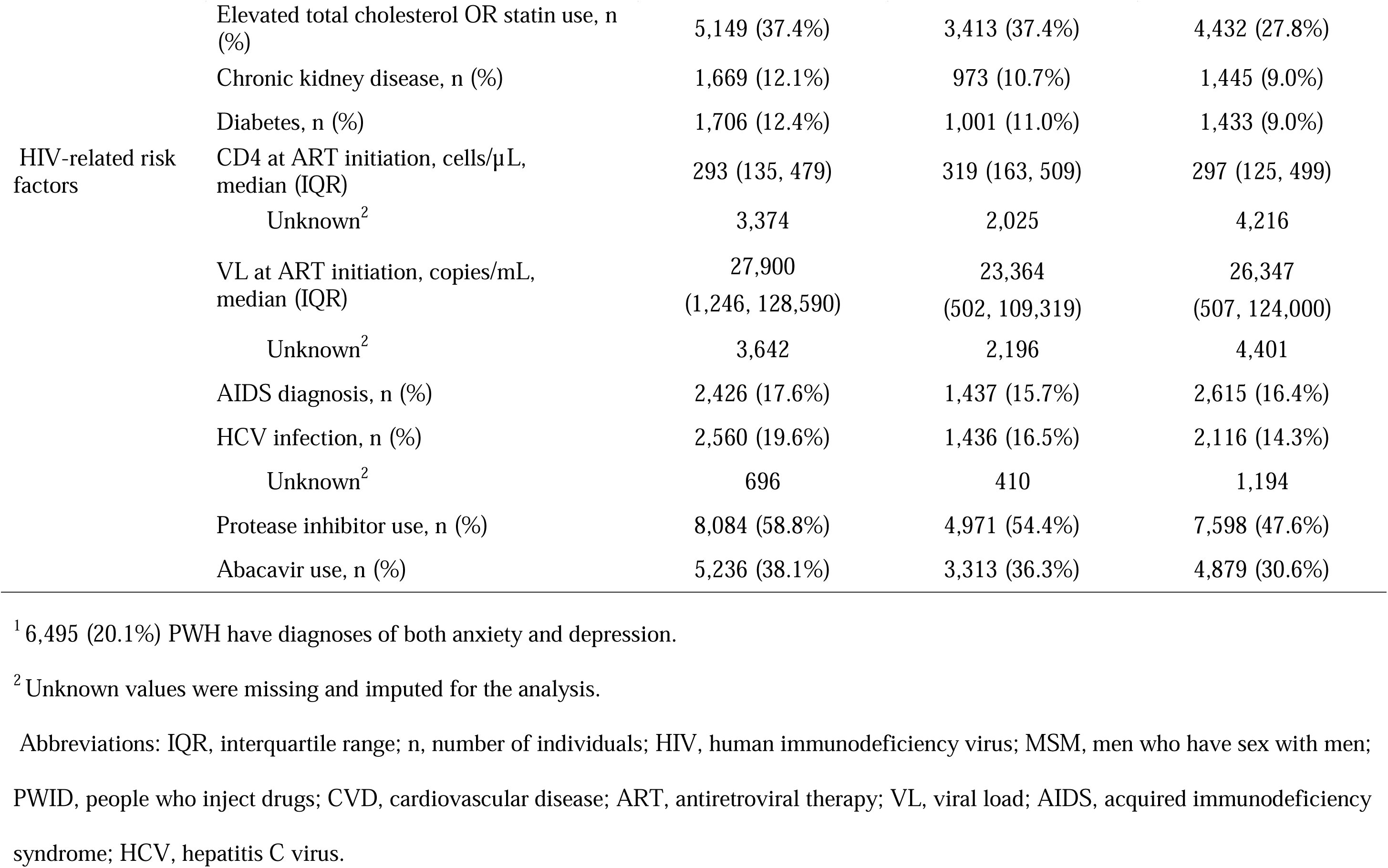
Demographic characteristics of participants, by anxiety, ever depression, or neither anxiety nor depression (N=32,358)

A larger proportion of PWH with a depression or anxiety diagnosis were non-Hispanic White, were PWID, had a history of substance use, and had traditional CVD risk factors (Table 1). People with a depression or anxiety diagnosis were more likely to have HCV co-infection, ever have a detectable VL after ART initiation, and ever have protease inhibitor or abacavir use. Compared with PWH with a depression diagnosis, PWH with an anxiety diagnosis were more likely to be non-Hispanic White, MSM, and have a higher CD4 and lower VL at ART initiation (Table 1).

Incident T1MI was diagnosed in 523 participants of whom 272 had depression and 148 had anxiety. T2MI occurred in 363 participants of whom 175 had depression and 95 had anxiety. Supplementary Figure 3 displays the median time prior to study entry or incident MI, depending on when the diagnosis occurred.

### The relationship of depression with T1MI and T2MI

PWH with (vs. without) a depression diagnosis had a greater cumulative incidence of T1MI over 20 years (9.4%, 95% CI [7.2-11.6%] vs. 7.8%, 95% CI [4.7, 10.8%], log rank p=<0.001, Figure 1A) but not an increased incidence of T2MI (5.0%, 95% CI [3.9-6.2%] vs. 5.0%, 95% CI [3.1, 6.8%], log rank p=0.07, Figure 1C).

**Figure 1.**
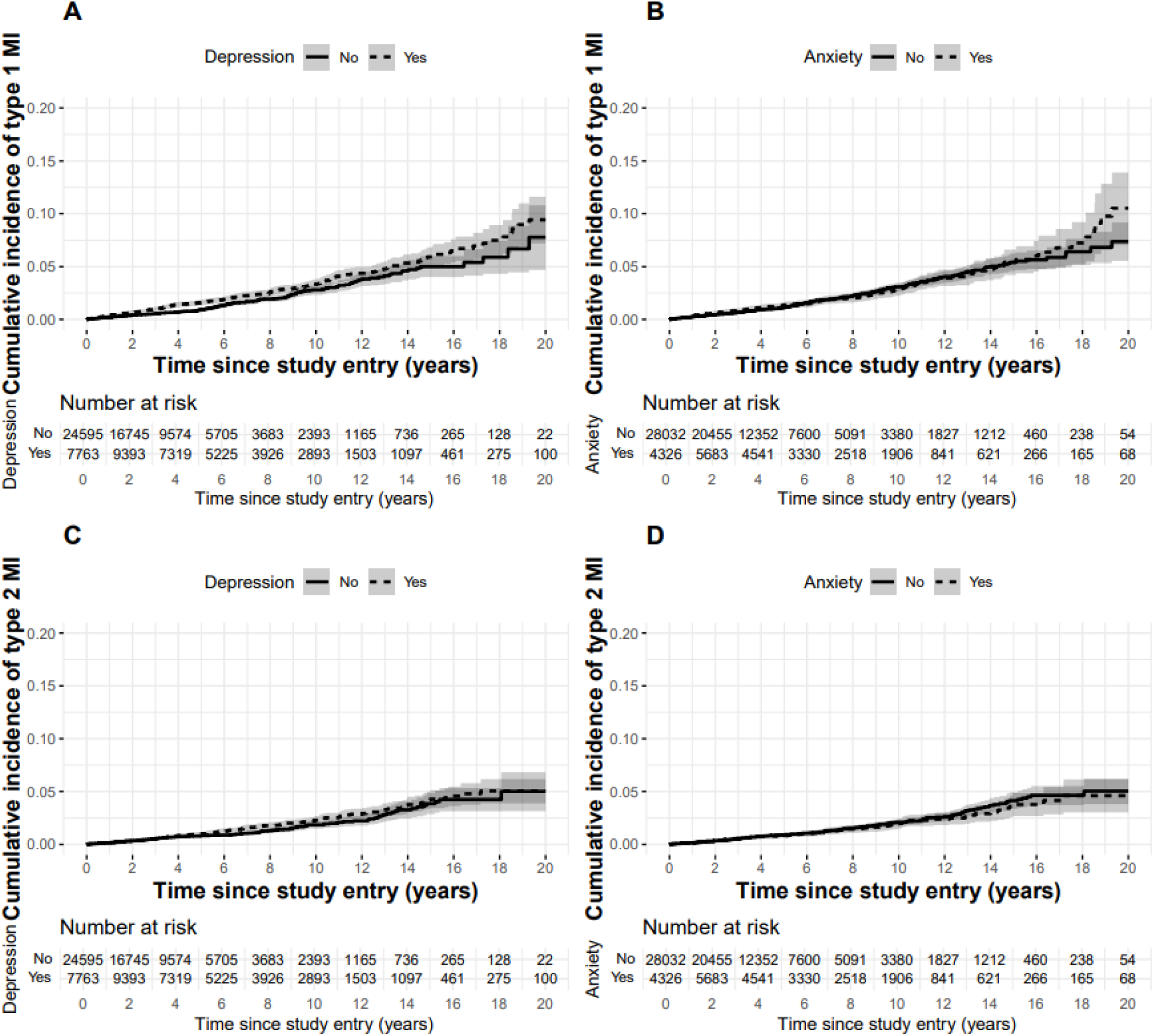
Kaplan-Meier estimates of the cumulative incidence of T1MI or T2MI, by depression and anxiety diagnosis. This figure displays the Kaplan-Meier estimates of the cumulative incidence of T1MI among people with vs without a depression diagnosis (Panel A) or with or without an anxiety diagnosis (Panel B), as well as the Kaplan-Meier estimates of the cumulative incidence of T2MI among people with vs without a depression diagnosis (Panel C) or with or without an anxiety diagnosis (Panel D).

In the unadjusted analysis, a depression diagnosis was strongly associated with incident T1MI, increasing T1MI risk by 38% during the study period (HR=1.38, 95% CI [1.15-1.64]) (Table 2). Controlling for demographics, anxiety, traditional CVD and HIV-related risk factors attenuated the association between depression diagnosis and T1MI risk, reducing the increased risk to 22% in PWH with (vs. without) a depression diagnosis (aHR=1.22, 95% CI [1.00-1.48]). Diagnosed depression was not associated with T2MI (aHR=1.05, 95% CI [0.83-1.33], Table 3).

**Table 2.** Estimated crude and adjusted hazard ratios and 95% confidence intervals from models of the risk of type 1 myocardial infarction.

| Characteristic | Unadjusted |  | Adjusted |  | Nested model including interaction term |  |
| --- | --- | --- | --- | --- | --- | --- |
|  | HR | 95% CI | aHR | 95% CI | aHR | 95% CI |
| Depression | 1.38 | 1.15, 1.64 | 1.22 | 1.00, 1.48 |  |  |
| Anxiety | 1.04 | 0.86, 1.26 | 0.86 | 0.70, 1.07 |  |  |
| Depression and/or anxiety |  |  |  |  |  |  |
| No depression no anxiety |  |  |  |  | 1.00 | -- |
| Yes depression no anxiety |  |  |  |  | 1.20 | 0.96, 1.49 |
| No depression yes anxiety |  |  |  |  | 0.81 | 0.55, 1.21 |
| Yes depression yes anxiety |  |  |  |  | 1.06 | 0.82, 1.36 |
| Age (scaled by 10) | 1.94 | 1.20, 1.99 | 1.50 | 1.35, 1.66 | 1.50 | 1.35, 1.66 |
| Male sex at birth | 1.55 | 1.20, 1.99 | 1.44 | 1.05, 1.97 | 1.44 | 1.5, 1.97 |
| Race/ethnicity |  |  |  |  |  |  |
| Non-Hispanic White | 1.00 | — | 1.00 | — | 1.00 | — |
| Non-Hispanic Black | 0.76 | 0.62, 0.92 | 0.68 | 0.53, 0.87 | 0.68 | 0.53, 0.87 |
| Hispanic | 0.55 | 0.40, 0.74 | 0.75 | 0.55, 1.03 | 0.75 | 0.55, 1.03 |
| Other/unknown | 0.80 | 0.55, 1.17 | 0.91 | 0.60, 1.39 | 0.91 | 0.60, 1.39 |
| HIV transmission risk group |  |  |  |  |  |  |
| Heterosexual contact | 1.00 | — | 1.00 | — | 1.00 | — |
| MSM | 1.26 | 1.02, 1.56 | 1.15 | 0.88, 1.50 | 1.15 | 0.88, 1.50 |
| PWID | 1.54 | 1.16, 2.04 | 1.18 | 0.84, 1.67 | 1.19 | 0.84, 1.67 |
| Other/unknown | 1.16 | 0.76, 1.79 | 0.81 | 0.50, 1.30 | 0.81 | 0.50, 1.30 |

| Characteristic | Unadjusted |  | Adjusted |  | Nested model including interaction term |  |
| --- | --- | --- | --- | --- | --- | --- |
|  | HR | 95% CI | aHR | 95% CI | aHR | 95% CI |
| Smoking | 2.31 | 1.83, 2.93 | 1.97 | 1.54, 2.53 | 1.97 | 1.54, 2.53 |
| At-risk alcohol use | 1.10 | 0.93, 1.32 | 1.00 | 0.82, 1.22 | 1.00 | 0.82, 1.22 |
| Cocaine use | 1.07 | 0.83, 1.39 | 0.97 | 0.71, 1.31 | 0.97 | 0.71, 1.31 |
| Cannabis use | 1.25 | 0.95, 1.63 | 1.22 | 0.91, 1.64 | 1.22 | 0.91, 1.64 |
| Hypertension | 5.15 | 4.24, 6.26 | 3.04 | 2.44, 3.78 | 3.04 | 2.44, 3.78 |
| Elevated total cholesterol or statin use | 3.86 | 3.19, 4.66 | 2.31 | 1.88, 2.85 | 2.31 | 1.88, 2.85 |
| Chronic kidney disease | 2.95 | 2.40, 3.62 | 1.23 | 0.98, 1.54 | 1.23 | 0.98, 1.55 |
| Diabetes | 2.63 | 2.14, 3.24 | 1.27 | 1.01, 1.59 | 1.27 | 1.01, 1.59 |
| AIDS diagnosis | 1.33 | 1.09, 1.62 | 0.99 | 0.78, 1.26 | 0.99 | 0.78, 1.26 |
| HCV infection | 1.33 | 1.08, 1.64 | 1.08 | 0.83, 1.39 | 1.08 | 0.82, 1.40 |
| Detectable viral load | 1.10 | 0.87, 1.40 | 1.25 | 0.95, 1.66 | 1.26 | 0.95, 1.66 |
| Protease inhibitor use | 1.76 | 1.48, 2.10 | 1.59 | 1.32, 1.91 | 1.59 | 1.32, 1.91 |
| Abacavir use | 1.73 | 1.43, 2.10 | 1.22 | 1.00, 1.50 | 1.22 | 1.00, 1.50 |

Model is also adjusted for CD4 count (time-varying), BMI, CD4 count at ART initiation, VL at ART initiation, and cohort. Abbreviations: MI, Myocardial Infarction; HR, Hazard Ratio; CI, Confidence Interval; MSM, men who have sex with men; PWID, people who inject drugs; AIDS, acquired immunodeficiency syndrome; HCV, hepatitis C virus; VL, viral load.

**Table 3.**
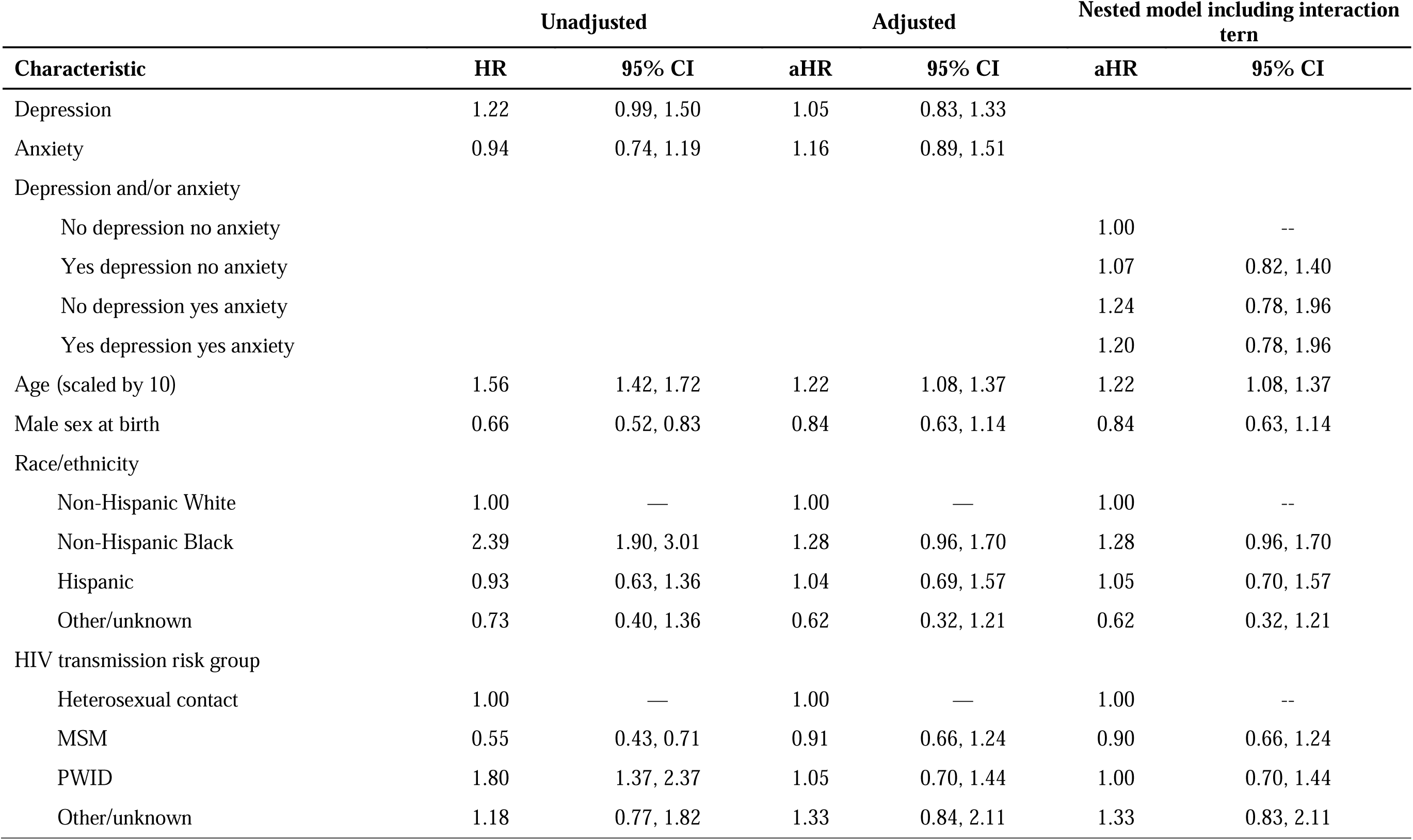
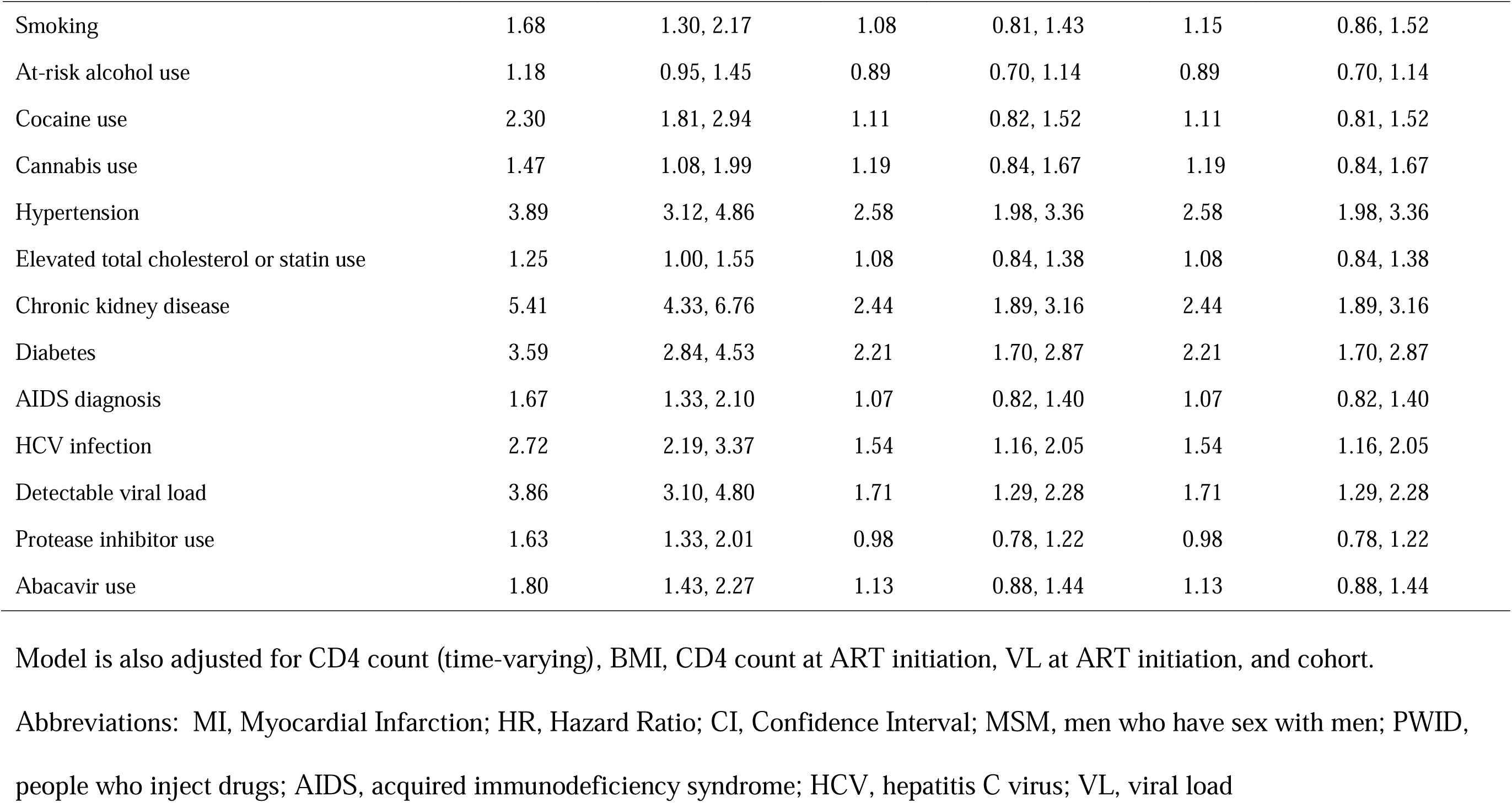
Estimated crude and adjusted hazard ratios and 95% confidence intervals from models of the risk of type 2 myocardial infarction.

### The relationship of anxiety with T1MI and T2MI

PWH with (vs. without) an anxiety diagnoses had a non-significantly increased cumulative incidence of T1MI over 20 years (10.5%, 95% CI [7.0-13.9%] vs. 7.4%, 95% CI [5.5-9.2%], log rank p=0.7, Figure 1B) and no increased cumulative incidence of T2MI (4.6%, 95% CI [3.0 - 6.2%] vs. 5.0%, 95% CI [3.8-6.2%], log rank p=0.6, Figure 1D).

The relationship between an anxiety diagnosis and T1MI or T2MI was not significant in the univariable or adjusted models. However, the adjusted model showed a protective association with T1MI (aHR=0.86, 95% CI [0.70-1.07], Table 2) and an increased association with T2MI (aHR=1.16, 95% CI [0.89-1.51], Table 3), although neither relationship was statistically significant.

### The relationship of depression with T1MI in PWH with and without anxiety

In the nested models, the model with the interaction for depression and anxiety did not have a better fit than the model without the interaction (p-value for T1MI models, 0.733; for T2MI models, 0.719).

Although not significant, we found that PWH with depression only had a higher risk of T1MI (aHR=1.20, 95% CI [0.96, 1.49]) compared to PWH without depression or anxiety, whereas PWH with anxiety only had a reduced association with T1MI (aHR=0.81, 95% CI [0.55, 1.21]. These opposing directions contributed to the null association observed among people with both depression *and* anxiety and T1MI in the nested model (aHR=1.06, 95% CI [0.82, 1.36]) (Table 2). Similarly, in models that included terms for depression or anxiety, without an interaction term, the association of depression or anxiety with T1MI was attenuated compared with when both variables were included in the model (Supplementary Table S4).

### The relationship of depression with T2MI in PWH with and without anxiety

The relationships between depression and anxiety and T2MI are presented in Table 3. Compared to PWH without depression or anxiety, PWH with depression only had a null association with T2MI (aHR=1.07, 95% CI [0.82, 1.40]), whereas PWH with anxiety only showed a non-significantly increased association with T2MI (aHR=1.24, 95% CI [0.78, 1.96]). Therefore, an increase of substantial magnitude was evident in the association of T2MI among PWH with both depression and anxiety, driven by anxiety (aHR=1.20, 95% CI [0.88, 1.65]), although the association was not statistically significant. Results from the models with only depression or anxiety were not substantially changed from the fully adjusted model (Supplementary Table S5).

### Sex-stratified results

Depression was associated with a 79% increased risk of T1MI among females (HR=1.79, 95% CI [1.08-2.96]), which was only slightly attenuated after adjustment for covariates but was no longer statistically significant (aHR=1.78, 95% CI [0.98-3.26]) (Supplementary Table 6). Among males, there was a 35% increased risk of T1MI among people with (vs without) a depression diagnosis (HR=1.35, 95% CI [1.12-1.63]), which attenuated and was no longer statistically significant in the fully adjusted model (aHR 1.16, 95% CI [0.94-1.43]).

Sex-stratified analyses for T2MI are shown in Supplementary Table 7.

## DISCUSSION

In this analysis of more than 30,000 people living with HIV in the US, we found a statistically significant 22% increase in the risk of T1MI among people with HIV who had a depression diagnosis compared to PWH without a diagnosis of depression. The magnitude of the association between depression diagnosis and T1MI underscores the need for mental health screening to identify PWH who are at increased risk for T1MI so that CVD primary prevention can be prioritized.

Depression has previously been associated with an increased risk of cardiovascular disease among people without HIV [7,12,13]. Depression is linked to autonomic nervous system dysregulation, characterized by increased sympathetic activity and reduced parasympathetic tone, which may predispose individuals to MI [23]. Depression also promotes chronic, low-grade systemic inflammation, with elevated pro-inflammatory cytokines that accelerate atherosclerosis and increase the risk of plaque rupture [24,25]. Neuroendocrine alterations, such as hyperactivity of the hypothalamic-pituitary-adrenal (HPA) axis and elevated cortisol levels), contribute to metabolic disturbances including central obesity, insulin resistance, and hypertension [26–28]. Thus, the co-occurrence of HIV and depression could synergistically increase the risk of T1MI through an interplay of these biological pathways, as well as behavioral factors like inconsistent medication adherence, substance use, and barriers to lifestyle-promoting activities. Together, these mechanisms may create a cumulative risk for T1MI with depression for PWH that is greater than the sum of their individual effects.

Our findings of no association of T2MI among PWH with (vs. without) diagnosed depression add to the growing understanding of T2MI in PWH. T2MI is caused by an imbalance between myocardial oxygen supply and demand, which can result from conditions like sepsis, substance use, or severe anemia [29]. Precipitants for T2MI are often acute and situational rather than driven by the chronic processes of inflammation and atherosclerosis as evident in T1MIs. These findings support previously published research from the general population that people with diagnosed depression have an increased risk for T1MI but not T2MI [30] and provide new evidence that this could be true for PWH as well.

Depression is a complex condition with major differences between sexes, including biologic and behavioral conditions. Notably, females without HIV diagnosed with depression have a relatively greater increased risk of cardiovascular outcomes than males, in particular at younger ages [31,32]. We found that the association between depression and T1MI among females had a magnitude of effect that was higher than evident with tobacco use or diabetes – and almost twice the effect seen among males with HIV, although it was not statistically significant in fully adjusted analysis. Hormonal differences may play a role in modulating the relationship between depression and cardiovascular outcomes; menopause, pregnancy, and higher rates of depression and anxiety in females have been correlated to higher prevalence of adverse cardiovascular outcomes [33,34]. Additionally, females exhibit a more robust inflammatory immune response in treatment of HIV [35], which could lead to a larger increase in risk of cardiovascular disease among females than males with HIV.

The association between diagnosed depression and incident T1MI was attenuated in sensitivity analysis that examined a shorter calendar timeframe after routine mental health screening was implemented in one cohort. Participants diagnosed with mood disorders later in the study period may have had less follow-up time, affecting the observed associations. Different screening tests and criteria for diagnosis of depression and anxiety could have influenced the timing of the diagnoses. People diagnosed with depression or anxiety prior to the implementation of routine screening could have more severe disease, which might increase their cardiovascular risk [36], given that the severity of depression has previously been reported to affect cardiovascular risk and outcomes [37–40] and duration of untreated depression has been associated with symptom severity and the likelihood of improvement over time [41]. Advances in HIV management and cardiovascular care since 2010 could also have mitigated some risks associated with depression and anxiety, potentially altering the observed relationships between these conditions and incident MI.

Although diagnosed anxiety was not significantly associated with incident T1MI or T2MI among PWH, we found non-significant but clinically substantial associations among PWH with diagnosed anxiety with each type of MI in the nested models: a 19% reduced risk of T1MI and a 24% increased risk of T2MI among people with anxiety only (i.e., without depression) compared to people without depression or anxiety. These findings suggest that the impact of distinct mental health comorbidities with each type of MI may be qualitatively different and may affect MI incidence through different mechanisms. In particular, the heterogeneous causes of T2MI (e.g., vasospasm, sepsis) may require a more detailed categorization of T2MI to better understand what demographics and other clinical exposures are associated with incident T2MI, such as substance use. Additionally, anxiety disorders are often less consistently documented by clinicians compared with depression, leading to under-diagnosis and limited documentation [42]; specific screening criteria for both anxiety and depression that can be applied uniformly and consistently are likely to help further elucidate associations with each type of MI event.

This study has several limitations. Although randomized controlled trials are the gold standard for assessing causation, an observational study is required to assess depression and anxiety with T1MI and T2MI because randomization to a depression or anxiety diagnosis is not possible. In an observational study, not accounting for all confounders is a threat to causality. We directly adjusted for known confounders and did not use inverse probability weighting to adjust for potential mediators given uncertainty in some data elements, such as the timing of when depression was diagnosed compared to its onset. Additionally, even if the associations presented in this study approximate the effect of depression on MI risk, that is not equivalent to the reduction in MI risk that might be realized by increasing treatment of prevalent depression [43]. The impact of treatment for, and management of, depression and/or anxiety was not evaluated due to a lack of data regarding specific psychiatric treatment within the study population and given medications for depression or anxiety can be used for treatment of multiple other diseases and are not specific to these mental health disorders. The reliance on ICD-coded diagnoses for anxiety and depression could result in misclassification and under-diagnosis.

In a large cohort study of PWH in the US, diagnosed depression was associated with T1MI; this relationship was stronger in females with HIV versus males with HIV. People with diagnosed anxiety had a non-significant but clinically substantial reduced association with T1MI and increased association with T2MI, suggesting that the impact of mental health comorbidities on type of MI is not the same. Further understanding of the role of mental health conditions, diagnosis, and treatment in improving cardiovascular health among PWH is needed.

## Data Availability

All data produced in the present study are available upon reasonable request to the authors

## NA-ACCORD Collaborating Cohorts and Representatives

<u>AIDS Clinical Trials Group Longitudinal Linked Randomized Trials</u>: Constance A. Benson and Ronald J. Bosch; <u>AIDS Link to the IntraVenous Experience</u>: Gregory D. Kirk; <u>Einstein-Rockefeller-CUNY CFAR Clinical Cohort</u>: Uriel R. Felsen and David B. Hanna <u>Emory-Grady HIV Clinical Cohort</u>: Vincent Marconi and Jonathan Colasanti; <u>Fenway Health HIV Cohort</u>: Kenneth H. Mayer; <u>HIV Outpatient Study</u>: Kate Buchacz and Jun Li; <u>HIV Research Network</u>: Kelly A. Gebo and Richard D. Moore; <u>Johns Hopkins HIV Clinical Cohort</u>: Richard D. Moore; John T. Carey Special Immunology Unit Patient Care and Research Database, Case Western Reserve University: George Yendewa; <u>Kaiser Permanente Mid-Atlantic States</u>: Michael A. Horberg; <u>Kaiser Permanente Northern California</u>: Michael J. Silverberg; <u>Longitudinal Study of Ocular Complications of AIDS</u>: Jennifer E. Thorne; <u>MACS/WIHS Combined Cohort Study</u>: Mirjam-Colette Kempf, Bradley Aouizerat, and Seble Kassaye; <u>Maple Leaf Medical Clinic</u>: Graham Smith, Mona Loutfy, and Meenakshi Gupta; <u>The McGill University Health Centre, Chronic Viral Illness Service Cohort</u>: Marina B. Klein; <u>Multicenter Hemophilia Cohort Study–II</u>: Charles Rabkin; <u>Parkland/UT Southwestern Cohort</u>: Ank Nijhawan; <u>Retrovirus Research Center, Universidad Central del Caribe, Bayamon Puerto Rico</u>: Angel M. Mayor; <u>Southern Alberta Clinic Cohort</u>: M. John Gill; <u>Study of the Consequences of the Protease Inhibitor Era</u>: Jeffrey N. Martin and Steven G. Deeks; <u>Study to Understand the Natural History of HIV/AIDS in the Era of Effective Therapy</u>: Kate Buchacz and Jun Li; <u>University of Alabama at Birmingham 1917 Clinic Cohort</u>: Michael S. Saag, Michael J. Mugavero, and Greer Burkholder; <u>University of California at San Diego</u>: Laura Bamford and Maile Karris; <u>University of North Carolina at Chapel Hill HIV Clinic Cohort</u>: Joseph J. Eron and Sonia Napravnik; <u>University of Washington HIV Cohort</u>: Mari M. Kitahata and Heidi M. Crane; <u>Vanderbilt Comprehensive Care Clinic HIV Cohort</u>: Timothy R. Sterling, David Haas, Peter Rebeiro, and Megan Turner; <u>Veterans Aging Cohort Study</u>: Lesley Park, Kathleen McGinnis, and Amy Justice

## NA-ACCORD Study Administration

<u>Executive Committee</u>: Keri N. Althoff, Richard D. Moore, Mari M. Kitahata, Catherine R. Lesko, Michael A. Horberg, Marina B. Klein, Rosemary G. McKaig, and Aimee M. Freeman; <u>Administrative Core</u>: Richard D. Moore, Keri N. Althoff, and Aimee M. Freeman; <u>Data Management Core</u>: Mari M. Kitahata, Heidi M. Crane, Joseph (Chris) Delaney, Liz Morton, Justin McReynolds, and William B. Lober; <u>Epidemiology and Biostatistics Core</u>: Catherine R. Lesko, Sally B. Coburn, Brenna Hogan, Elizabeth Humes, Wendy (Chunyan) Zheng, Jason Haw, Yawei Cheng, and Lucas Gerace.

## SUPPLEMENTARY APPENDIX

## Supplementary Methods

### Sensitivity Analysis

In sensitivity analysis, we restricted the observation window to 2010-2019, when a marked increase in depression and anxiety diagnoses was noted following a change in screening protocols for mental health disorders in one of the contributing cohorts.

## Supplementary Results

### Sensitivity Analysis

In the sensitivity analysis that restricted one cohort to an observation window of 2010-2019 (given changes in clinical practice of mental health screening), the overall cohort size decreased from 32,358 to 31,524 PWH, total person-years decreased from 176,310.3 to 167,284.6 with no major changes in demographic characteristics, and 495 T1MIs and 353 T2MIs occurred (Supplementary Table 8). The magnitude of the effect size in the association between depression and T1MI was slightly reduced and was no longer statistically significant (aHR=1.19, 95% CI [0.97-1.45], Supplementary Figure 4). All other covariates and associations were not substantively changed (Supplementary Figures 4 and 5).

## Supplementary Tables

**Supplementary Table 1.**
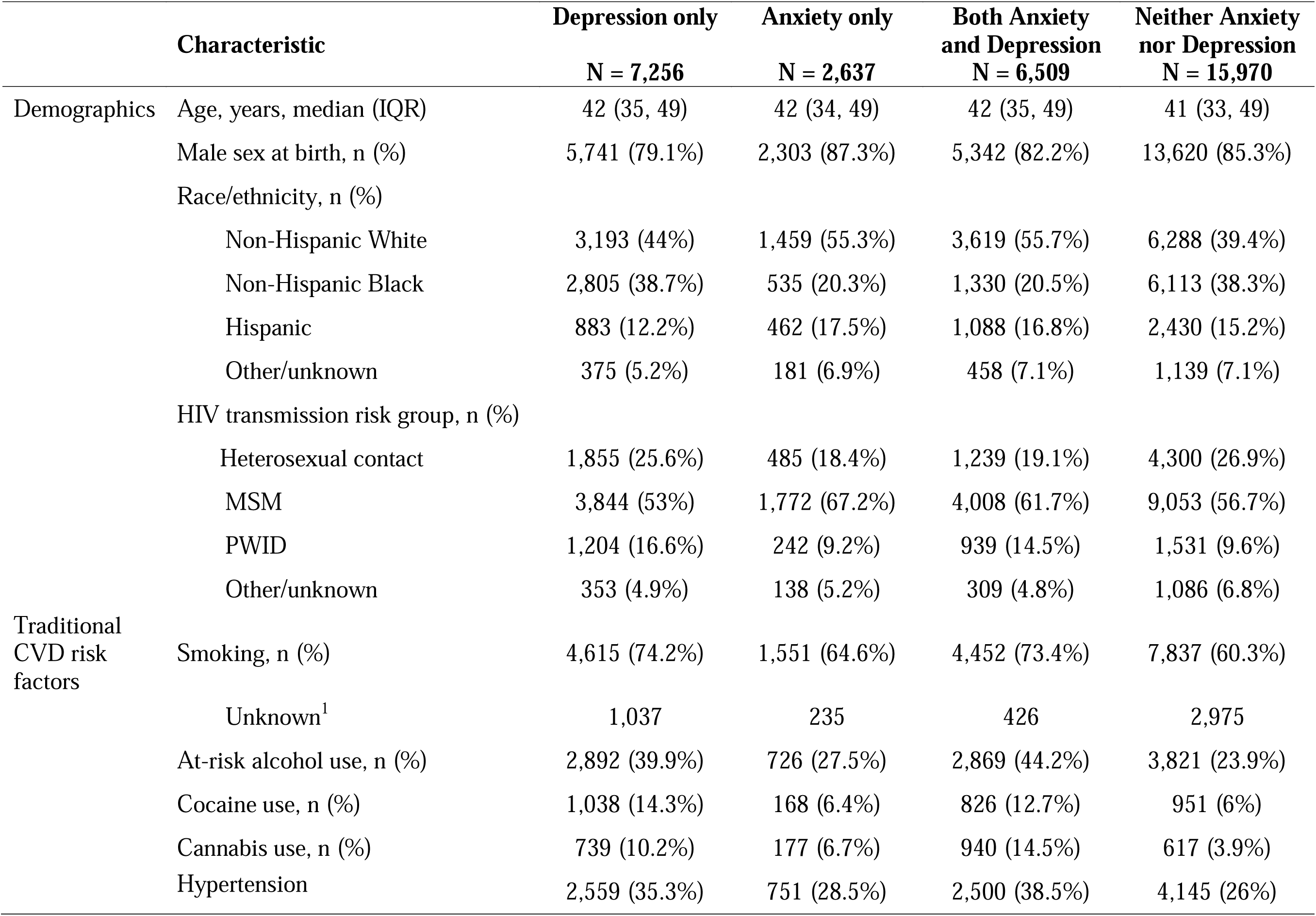
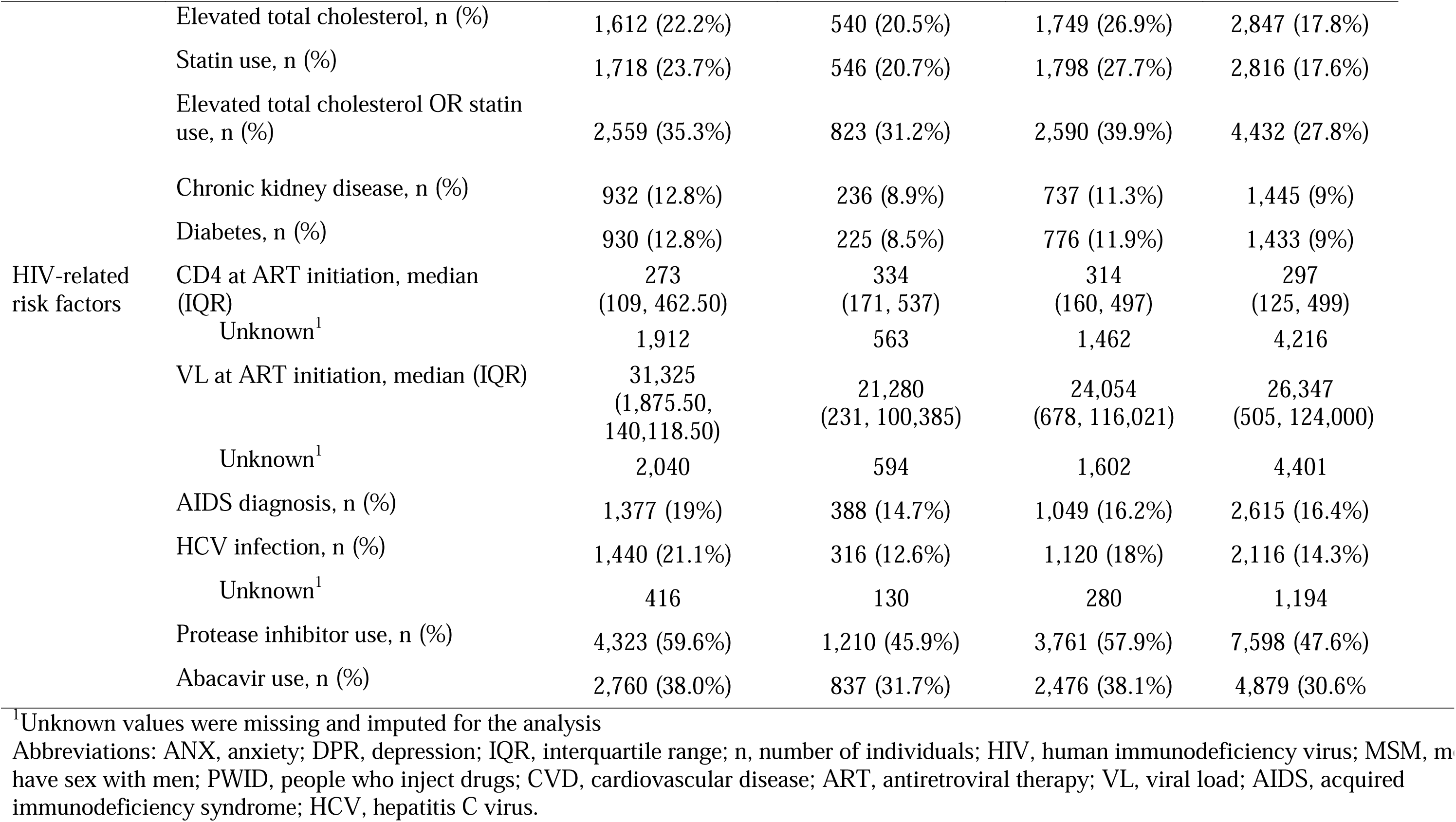
Demographic characteristics of participants by depression only, anxiety only, both anxiety and depression, or neither anxiety nor depression. (N=32,358)

**Supplementary Table 2:** Time-fixed variable definitions.

| **Variable** | **Definition** | **When assessed** | **Parameterization** |
| --- | --- | --- | --- |
| Sex at birth | Sex at birth | Enrollment into NA-ACCORD | Binary (male/female) |
| Age (scaled by 10) | Age at study entry | Study entry date | Continuous variable with a single linear term |
| Race/ethnicity | Race/ethnicity | Enrollment into NA-ACCORD | Categorical variable (Non-Hispanic white[ref], non-Hispanic Black, Hispanic, Other/unknown) |
| HIV acquisition risk group | HIV acquisition risk group | Enrollment into NA-ACCORD | Categorical variable (Heterosexual contact [ref], MSM, PWID, Other/unknown) |
| Cohort | Cohort membership | Enrollment into NA-ACCORD | Categorical |
| Smoking | Tobacco use | All data available at all time points | Binary (never/ever) |
| Alcohol misuse | Alcohol use/dependence defined as having an alcohol abuse or dependence diagnosis OR ever reporting at-risk alcohol use or binge drinking on a patient reported outcome (PRO) survey | All data available at all time points | Binary (never/ever) |
| Cocaine abuse/dependence | Cocaine abuse or dependence diagnosis | All data available at all time points | Binary (never/ever) |

**Supplementary Table 2: Time-fixed variable definitions (Con't)**
| Variable | Definition | When assessed | Parameterization |
| --- | --- | --- | --- |
| Cannabis abuse/dependence | Cannabis abuse or dependence diagnosis | All data available at all time points | Binary (never/ever) |
| HCV | Patient ever had evidence of HCV infection while under observation, defined as 1) a positive HCV antibody test, 2) a detectable HCV RNA, or 3) the presence of an HCV genotype test | All data available at all time points | Binary (no/yes) |
| CD4 at ART initiation | CD4 at ART initiation (closest lab measurement to date of ART initiation in the window of 1 year prior to ART initiation through 30 days after) | ART initiation | 4-knot restricted cubic spline |
| VL at ART initiation | VL at ART initiation (closest lab measurement to date of ART initiation in the window of 1 year prior to ART initiation through 30 days after) | ART initiation | 4-knot restricted cubic spline |
| AIDS at ART initiation | Patient had an AIDS-defining illness on or prior to ART initiation date | ART initiation | Binary (no/yes) |

Abbreviations: HIV, human immunodeficiency virus; HCV, hepatitis C virus; ART, antiretroviral therapy; VL, viral load; AIDS, acquired immunodeficiency syndrome.

**Supplementary Table 3:**
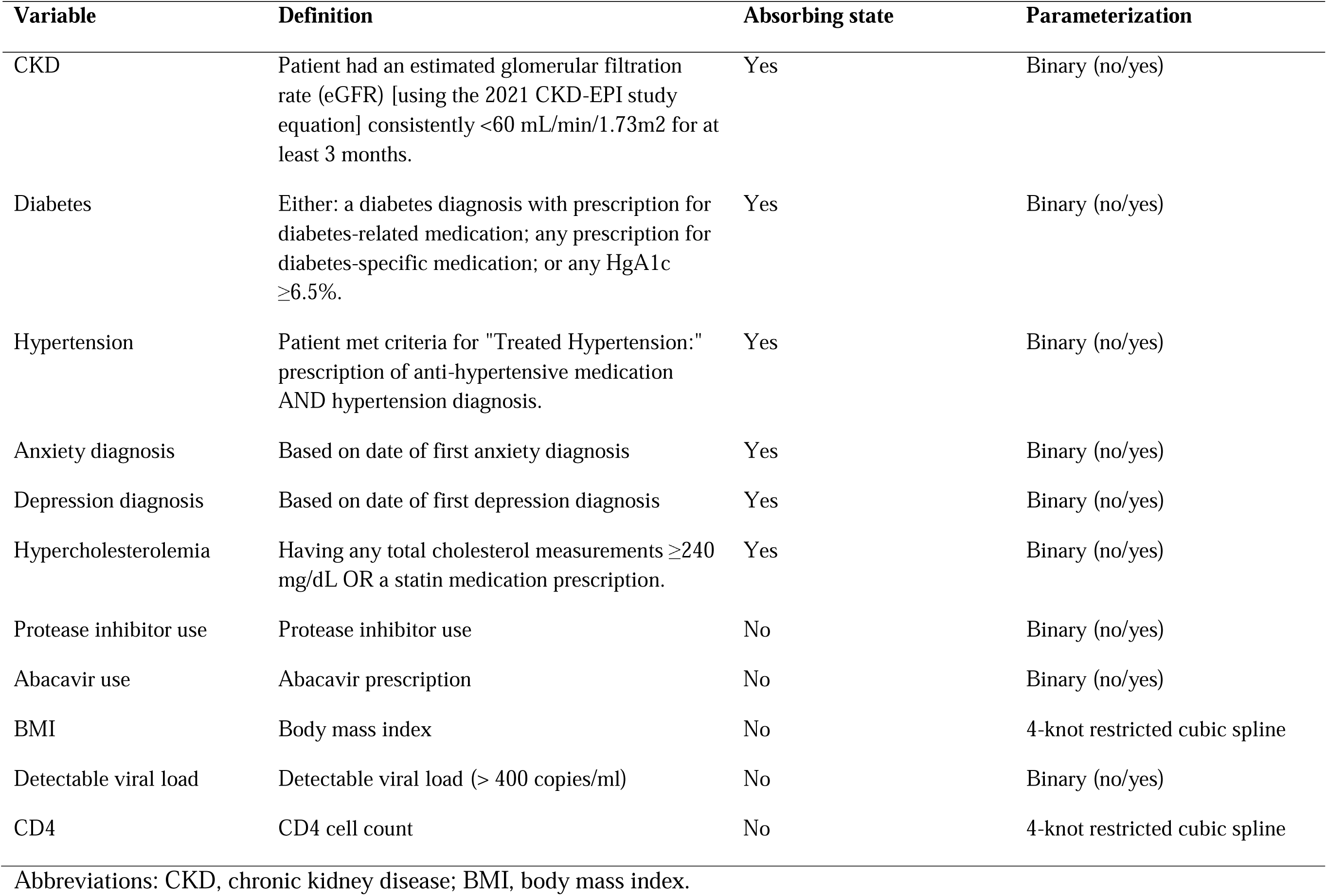
Time-varying variable definitions.

| Variable | Definition | Absorbing state | Parameterization |
| --- | --- | --- | --- |
| CKD | Patient had an estimated glomerular filtration rate (eGFR) [using the 2021 CKD-EPI study equation] consistently <60 mL/min/1.73m<sup>2</sup> for at least 3 months. | Yes | Binary (no/yes) |
| Diabetes | Either: a diabetes diagnosis with prescription for diabetes-related medication; any prescription for diabetes-specific medication; or any HgA1c ≥6.5%. | Yes | Binary (no/yes) |
| Hypertension | Patient met criteria for "Treated Hypertension:" prescription of anti-hypertensive medication AND hypertension diagnosis. | Yes | Binary (no/yes) |
| Anxiety diagnosis | Based on date of first anxiety diagnosis | Yes | Binary (no/yes) |
| Depression diagnosis | Based on date of first depression diagnosis | Yes | Binary (no/yes) |
| Hypercholesterolemia | Having any total cholesterol measurements ≥240 mg/dL OR a statin medication prescription. | Yes | Binary (no/yes) |
| Protease inhibitor use | Protease inhibitor use | No | Binary (no/yes) |
| Abacavir use | Abacavir prescription | No | Binary (no/yes) |
| BMI | Body mass index | No | 4-knot restricted cubic spline |
| Detectable viral load | Detectable viral load (> 400 copies/ml) | No | Binary (no/yes) |
| CD4 | CD4 cell count | No | 4-knot restricted cubic spline |

Abbreviations: CKD, chronic kidney disease; BMI, body mass index.

**Supplementary Table 4:**
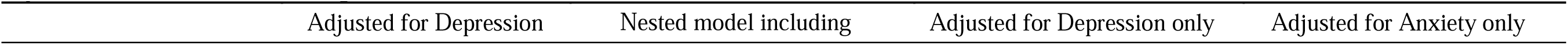
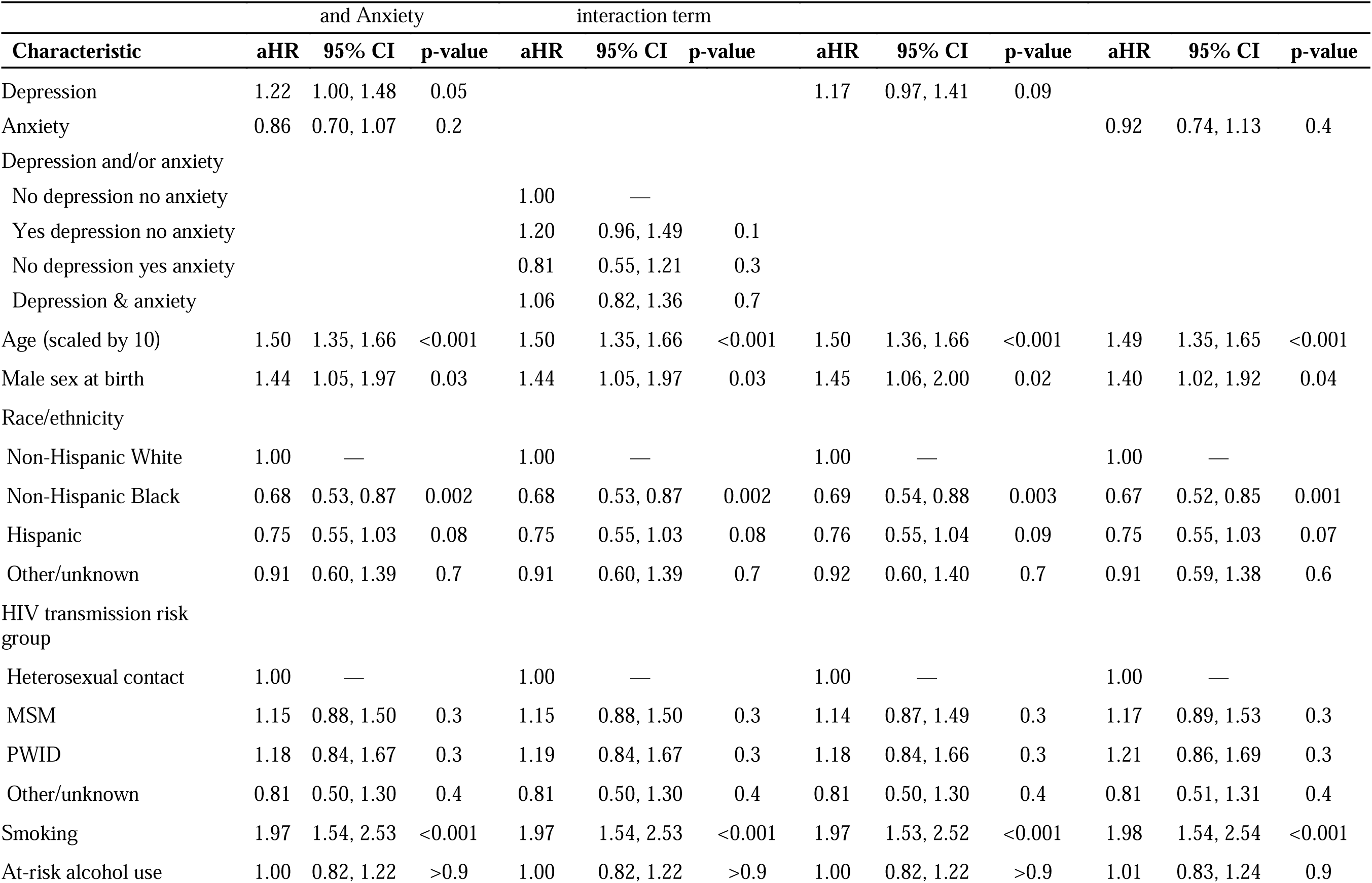
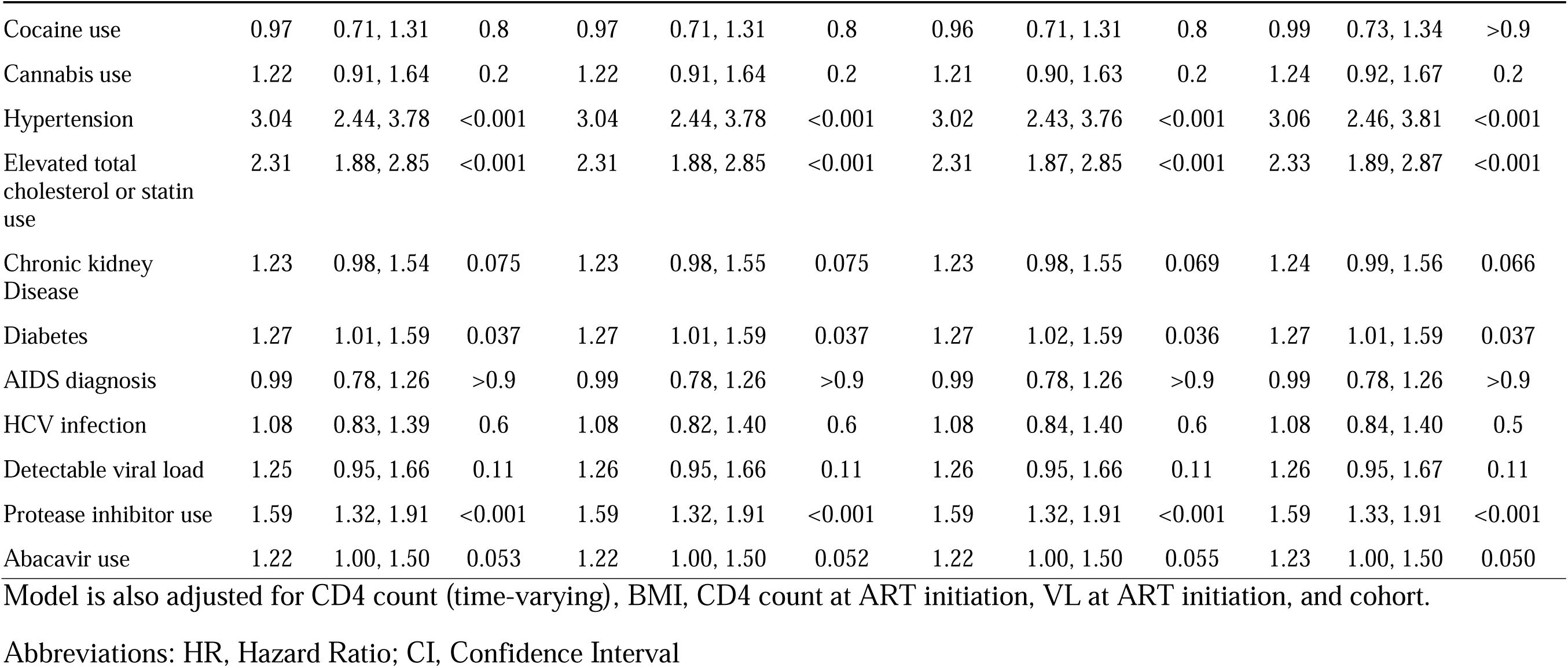
Estimated adjusted hazard ratios and 95% confidence intervals from models of the risk of type 1 myocardial infarction, using four different models.

**Supplementary Table 5:** Estimated adjusted hazard ratios and 95% confidence intervals from models of the risk of type 2 myocardial infarction, using four different models.

| Characteristic | Adjusted for Depression and Anxiety |  |  | Nested model including interaction term |  |  | Adjusted for Depression only |  |  | Adjusted for Anxiety only |  |  |
| --- | --- | --- | --- | --- | --- | --- | --- | --- | --- | --- | --- | --- |
|  | aHR | 95% CI | p-value | aHR | 95% CI | p-value | aHR | 95% CI | p-value | aHR | 95% CI | p-value |
| Depression | 1.05 | 0.83, 1.33 | 0.7 |  |  |  | 1.08 | 0.86, 1.37 | 0.5 |  |  |  |
| Anxiety | 1.16 | 0.89, 1.51 | 0.3 |  |  |  |  |  |  | 1.17 | 0.91, 1.52 | 0.2 |
| Depression and/or anxiety |  |  |  |  |  |  |  |  |  |  |  |  |
| No depression no anxiety |  |  |  | 1.00 | — |  |  |  |  |  |  |  |
| Yes depression no anxiety |  |  |  | 1.07 | 0.82, 1.40 | 0.4 |  |  |  |  |  |  |
| No depression yes anxiety |  |  |  | 1.24 | 0.78, 1.96 | 0.6 |  |  |  |  |  |  |
| Depression & anxiety |  |  |  | 1.20 | 0.78, 1.96 | 0.3 |  |  |  |  |  |  |
| Age (scaled by 10) | 1.22 | 1.08, 1.37 | 0.001 | 1.22 | 1.08, 1.37 | 0.001 | 1.22 | 1.08, 1.37 | 0.001 | 1.22 | 1.08, 1.37 | 0.001 |
| Male sex at birth | 0.84 | 0.63, 1.14 | 0.3 | 0.84 | 0.63, 1.14 | 0.3 | 0.83 | 0.62, 1.12 | 0.2 | 0.84 | 0.62, 1.12 | 0.2 |
| Race/ethnicity |  |  |  |  |  |  |  |  |  |  |  |  |
| Non-Hispanic White | 1.00 | — |  | 1.00 | — |  | 1.00 | — |  | 1.00 | — |  |
| Non-Hispanic Black | 1.28 | 0.96, 1.70 | 0.09 | 1.28 | 0.96, 1.70 | 0.09 | 1.26 | 0.95, 1.68 | 0.1 | 1.28 | 0.96, 1.70 | 0.1 |
| Hispanic | 1.04 | 0.69, 1.57 | 0.8 | 1.05 | 0.70, 1.57 | 0.8 | 1.04 | 0.69, 1.56 | 0.9 | 1.04 | 0.69, 1.57 | 0.8 |
| Other/unknown | 0.62 | 0.32, 1.21 | 0.2 | 0.62 | 0.32, 1.21 | 0.2 | 0.61 | 0.31, 1.18 | 0.1 | 0.62 | 0.32, 1.21 | 0.2 |
| HIV transmission risk group |  |  |  |  |  |  |  |  |  |  |  |  |
| Heterosexual contact | 1.00 | — |  | 1.00 | — |  | 1.00 | — |  | 1.00 | — |  |
| MSM | 0.91 | 0.66, 1.24 | 0.5 | 0.90 | 0.66, 1.24 | 0.5 | 0.91 | 0.66, 1.25 | 0.6 | 0.91 | 0.66, 1.25 | 0.6 |

| Characteristic | Adjusted for Depression and Anxiety |  |  | Nested model including interaction term |  |  | Adjusted for Depression only |  |  | Adjusted for Anxiety only |  |  |
| --- | --- | --- | --- | --- | --- | --- | --- | --- | --- | --- | --- | --- |
|  | aHR | 95% CI | p-value | aHR | 95% CI | p-value | aHR | 95% CI | p-value | aHR | 95% CI | p-value |
| PWID | 1.00 | 0.70, 1.44 | >0.9 | 1.00 | 0.70, 1.44 | >0.9 | 1.01 | 0.70, 1.44 | >0.9 | 1.01 | 0.70, 1.45 | >0.9 |
| Other/unknown | 1.33 | 0.84, 2.11 | 0.2 | 1.33 | 0.83, 2.11 | 0.2 | 1.33 | 0.84, 2.12 | 0.2 | 1.33 | 0.84, 2.11 | 0.2 |
| Smoking | 1.15 | 0.86, 1.52 | 0.3 | 1.15 | 0.86, 1.52 | 0.3 | 1.15 | 0.86, 1.53 | 0.3 | 1.15 | 0.86, 1.52 | 0.3 |
| At-risk alcohol use | 0.89 | 0.70, 1.14 | 0.4 | 0.89 | 0.70, 1.14 | 0.4 | 0.90 | 0.70, 1.14 | 0.4 | 0.90 | 0.70, 1.14 | 0.4 |
| Cocaine use | 1.11 | 0.82, 1.52 | 0.5 | 1.11 | 0.81, 1.52 | 0.5 | 1.12 | 0.82, 1.52 | 0.5 | 1.12 | 0.82, 1.52 | 0.5 |
| Cannabis use | 1.19 | 0.84, 1.67 | 0.3 | 1.19 | 0.84, 1.67 | 0.3 | 1.19 | 0.84, 1.67 | 0.3 | 1.19 | 0.85, 1.68 | 0.3 |
| Hypertension | 2.58 | 1.98, 3.36 | <0.001 | 2.58 | 1.98, 3.36 | <0.001 | 2.59 | 1.99, 3.38 | <0.001 | 2.58 | 1.98, 3.37 | <0.001 |
| Elevated total cholesterol or statin use | 1.08 | 0.84, 1.38 | 0.6 | 1.08 | 0.84, 1.38 | 0.6 | 1.08 | 0.84, 1.38 | 0.6 | 1.08 | 0.84, 1.38 | 0.6 |
| Chronic kidney disease | 2.44 | 1.89, 3.16 | <0.001 | 2.44 | 1.89, 3.16 | <0.001 | 2.44 | 1.88, 3.15 | <0.001 | 2.45 | 1.89, 3.17 | <0.001 |
| Diabetes | 2.21 | 1.70, 2.87 | <0.001 | 2.21 | 1.70, 2.87 | <0.001 | 2.21 | 1.70, 2.87 | <0.001 | 2.21 | 1.70, 2.87 | <0.001 |
| AIDS diagnosis | 1.07 | 0.82, 1.40 | 0.6 | 1.07 | 0.82, 1.40 | 0.6 | 1.07 | 0.82, 1.40 | 0.6 | 1.08 | 0.82, 1.41 | 0.6 |
| HCV infection | 1.54 | 1.16, 2.05 | 0.003 | 1.54 | 1.16, 2.05 | 0.003 | 1.54 | 1.16, 2.05 | 0.003 | 1.55 | 1.16, 2.05 | 0.003 |
| Detectable viral load | 1.71 | 1.29, 2.28 | <0.001 | 1.71 | 1.29, 2.28 | <0.001 | 1.71 | 1.29, 2.28 | <0.001 | 1.71 | 1.29, 2.28 | <0.001 |
| Protease inhibitor use | 0.98 | 0.78, 1.22 | 0.8 | 0.98 | 0.78, 1.22 | 0.8 | 0.98 | 0.78, 1.22 | 0.8 | 0.98 | 0.78, 1.22 | 0.8 |
| Abacavir use | 1.13 | 0.88, 1.44 | 0.3 | 1.13 | 0.88, 1.44 | 0.3 | 1.13 | 0.88, 1.45 | 0.3 | 1.13 | 0.88, 1.44 | 0.3 |

Model is also adjusted for CD4 count (time-varying), BMI, CD4 count at ART initiation, VL at ART initiation, and cohort.
Abbreviations: HR, Hazard Ratio; CI, Confidence Interval

**Supplementary Table 6.** Sex-stratified crude and adjusted hazard ratios and 95% confidence intervals from models of the risk of Type 1 myocardial infarction (MI).

| Characteristic | Male |  |  |  | Female |  |  |  |
| --- | --- | --- | --- | --- | --- | --- | --- | --- |
|  | Unadjusted |  | Adjusted |  | Unadjusted |  | Adjusted |  |
|  | HR<sup>1</sup> | 95% CI<sup>1</sup> | aHR | 95% CI | HR | 95% CI | aHR | 95% CI |
| Depression | 1.35 | 1.12, 1.63 | 1.16 | 0.94, 1.43 | 1.79 | 1.08, 2.96 | 1.78 | 0.98, 3.26 |
| Anxiety | 1.03 | 0.84, 1.27 | 0.86 | 0.68, 1.08 | 1.03 | 0.59, 1.79 | 0.94 | 0.49, 1.79 |
| Age (scaled by 10) | 1.96 | 1.80, 2.13 | 1.53 | 1.37, 1.71 | 1.85 | 1.51, 2.27 | 1.33 | 1.00, 1.76 |
| Race/ethnicity, |  |  |  |  |  |  |  |  |
| Non-Hispanic White | 1.00 | — | 1.00 | — | 1.00 | — | 1.00 | — |
| Non-Hispanic Black | 0.78 | 0.63, 0.97 | 0.67 | 0.51, 0.87 | 1.19 | 0.66, 2.13 | 0.83 | 0.41, 1.68 |
| Hispanic | 0.54 | 0.39, 0.74 | 0.73 | 0.52, 1.01 | 0.77 | 0.28, 2.12 | 1.20 | 0.39, 3.73 |
| Other/unknown | 0.80 | 0.53, 1.20 | 0.90 | 0.57, 1.40 | 1.23 | 0.41, 3.70 | 1.36 | 0.35, 5.33 |
| HIV transmission risk group |  |  |  |  |  |  |  |  |
| Heterosexual contact | 1.00 | — | 1.00 | — | 1.00 | — | 1.00 | — |
| MSM | 1.00 | 0.77, 1.31 | 1.15 | 0.86, 1.54 | — | — | — | — |
| PWID | 1.31 | 0.94, 1.83 | 1.25 | 0.85, 1.83 | 1.42 | 0.76, 2.66 | 0.76 | 0.31, 1.83 |
| Other/unknown | 0.97 | 0.59, 1.60 | 0.83 | 0.45, 1.32 | 1.23 | 0.49, 3.08 | 1.09 | 0.35, 3.36 |
| Smoking | 2.32 | 1.79, 3.00 | 2.02 | 1.52, 2.67 | 2.12 | 1.10, 4.06 | 1.67 | 0.77, 3.61 |
| At-risk alcohol use | 1.06 | 0.88, 1.28 | 1.00 | 0.81, 1.24 | 1.21 | 0.74, 2.00 | 1.03 | 0.54, 1.94 |

| Characteristic | Male |  |  |  | Female |  |  |  |
| --- | --- | --- | --- | --- | --- | --- | --- | --- |
|  | Unadjusted |  | Adjusted |  | Unadjusted |  | Adjusted |  |
|  | HR | 95% CI | aHR | 95% CI | HR | 95% CI | aHR | 95% CI |
| Cocaine use | 1.11 | 0.83, 1.47 | 1.05 | 0.75, 1.48 | 1.05 | 0.55, 2.00 | 0.53 | 0.22, 1.27 |
| Cannabis use | 1.19 | 0.89, 1.58 | 1.15 | 0.84, 1.58 | 1.58 | 0.76, 3.31 | 1.76 | 0.71, 4.36 |
| Hypertension | 5.14 | 4.18, 6.32 | 2.91 | 2.31, 3.67 | 6.04 | 3.38, 10.8 | 3.87 | 1.89, 7.93 |
| Elevated total cholesterol or statin use | 4.07 | 3.31, 4.99 | 2.41 | 1.92, 3.02 | 2.74 | 1.66, 4.51 | 1.72 | 0.94, 3.16 |
| Chronic kidney disease | 2.81 | 2.24, 3.53 | 1.16 | 0.90, 1.49 | 4.55 | 2.74, 7.53 | 1.87 | 1.01, 3.46 |
| Diabetes | 2.79 | 2.23, 3.50 | 1.27 | 1.00, 1.61 | 2.26 | 1.29, 3.94 | 1.36 | 0.70, 2.61 |
| AIDS diagnosis | 1.28 | 1.03, 1.58 | 0.97 | 0.75, 1.25 | 1.60 | 0.93, 2.74 | 1.19 | 0.58, 2.41 |
| HCV infection | 1.26 | 1.01, 1.59 | 1.04 | 0.79, 1.38 | 1.84 | 1.08, 3.14 | 1.54 | 0.74, 3.22 |
| Detectable viral load after ART initiation | 1.09 | 0.84, 1.42 | 1.27 | 0.93, 1.72 | 1.32 | 0.74, 2.34 | 1.15 | 0.53, 2.48 |
| Protease inhibitor use | 1.81 | 1.50, 2.18 | 1.57 | 1.29, 1.90 | 1.60 | 0.98, 2.62 | 1.78 | 1.00, 3.17 |
| Abacavir use | 1.65 | 1.33, 2.03 | 1.20 | 0.96, 1.49 | 2.41 | 1.46, 3.97 | 1.41 | 0.79, 2.52 |

Model is also adjusted for CD4 count (time-varying), BMI, CD4 count at ART initiation, VL at ART initiation, and cohort.
Abbreviations: HR, Hazard Ratio; CI, Confidence Interval

**Supplementary Table 7.** Sex-stratified crude and adjusted hazard ratios and 95% confidence intervals from models of the risk of Type 2 myocardial infarction (MI).

| Characteristic | Male |  |  |  | Female |  |  |  |
| --- | --- | --- | --- | --- | --- | --- | --- | --- |
|  | Unadjusted |  | Adjusted |  | Unadjusted |  | Adjusted |  |
|  | HR | 95% CI | aHR | 95% CI | HR | 95% CI | aHR | 95% CI |
| Depression | 1.18 | 0.92, 1.50 | 1.06 | 0.80, 1.40 | 1.24 | 0.82, 1.88 | 1.00 | 0.60, 1.67 |
| Anxiety | 0.94 | 0.71, 1.23 | 1.20 | 0.87, 1.64 | 0.97 | 0.60, 1.56 | 1.10 | 0.64, 1.89 |
| Age (scaled by 10) | 1.61 | 1.44, 1.81 | 1.20 | 1.04, 1.39 | 1.47 | 1.23, 1.76 | 1.30 | 1.04, 1.62 |
| Race/ethnicity, |  |  |  |  |  |  |  |  |
| Non-Hispanic White | 1.00 | — | 1.00 | — | 1.00 | — | 1.00 | — |
| Non-Hispanic Black | 2.44 | 1.87, 3.17 | 1.32 | 0.95, 1.84 | 1.78 | 1.03, 3.06 | 1.11 | 0.59, 2.10 |
| Hispanic | 0.93 | 0.61, 1.41 | 1.01 | 0.65, 1.58 | 0.84 | 0.33, 2.16 | 1.04 | 0.35, 3.10 |
| Other/unknown | 0.84 | 0.44, 1.61 | 0.75 | 0.35, 1.43 | 0.29 | 0.04, 2.16 | 0.31 | 0.04, 2.50 |
| HIV transmission risk group |  |  |  |  |  |  |  |  |
| Heterosexual contact | 1.00 | — | 1.00 | — | 1.00 | — | 1.00 | — |
| MSM | 0.49 | 0.36, 0.66 | 0.77 | 0.55, 1.08 | — | — | — | — |
| PWID | 1.44 | 1.02, 2.04 | 0.86 | 0.56, 1.32 | 2.79 | 1.75, 4.44 | 1.15 | 0.58, 2.26 |
| Other/unknown | 0.71 | 0.39, 1.29 | 0.75 | 0.40, 1.43 | 2.70 | 1.45, 5.04 | 3.32 | 1.67, 6.61 |
| Smoking | 1.79 | 1.32, 2.43 | 1.20 | 0.86, 1.68 | 1.52 | 0.93, 2.48 | 1.01 | 0.57, 1.80 |
| At-risk alcohol use | 1.14 | 0.90, 1.46 | 0.82 | 0.62, 1.09 | 1.44 | 0.95, 2.18 | 1.19 | 0.71, 2.01 |
| Cocaine use | 2.26 | 1.69, 3.03 | 1.18 | 0.82, 1.70 | 2.19 | 1.40, 3.43 | 0.89 | 0.47, 1.70 |
| Cannabis use | 1.29 | 0.89, 1.85 | 1.11 | 0.74, 1.67 | 2.26 | 1.30, 3.93 | 1.42 | 0.71, 2.83 |
| Hypertension | 4.24 | 3.28, 5.50 | 2.75 | 2.02, 3.76 | 2.83 | 1.84, 4.34 | 1.88 | 1.11, 3.17 |
| Elevated total cholesterol<br>or statin use | 1.29 | 1.00, 1.66 | 1.09 | 0.82, 1.46 | 1.13 | 0.74, 1.74 | 1.05 | 0.63, 1.74 |

| Characteristic | Male |  |  |  | Female |  |  |  |
| --- | --- | --- | --- | --- | --- | --- | --- | --- |
|  | Unadjusted |  | Adjusted |  | Unadjusted |  | Adjusted |  |
|  | HR | 95% CI | aHR | 95% CI | HR | 95% CI | aHR | 95% CI |
| Chronic kidney disease | 5.56 | 4.28, 7.22 | 2.30 | 1.70, 3.12 | 4.64 | 3.03, 7.10 | 2.86 | 1.72, 4.74 |
| Diabetes | 4.41 | 3.38, 5.75 | 2.55 | 1.89, 3.44 | 1.79 | 1.09, 2.96 | 1.44 | 0.81, 2.55 |
| AIDS diagnosis | 1.53 | 1.17, 2.00 | 1.05 | 0.77, 1.44 | 2.17 | 1.42, 3.34 | 1.14 | 0.66, 1.97 |
| HCV infection | 2.70 | 2.10, 3.47 | 1.48 | 1.07, 2.06 | 2.70 | 1.77, 4.12 | 1.76 | 0.98, 3.16 |
| Detectable viral load after ART initiation | 3.48 | 2.69, 4.52 | 1.75 | 1.25, 2.45 | 4.63 | 3.03, 7.06 | 1.66 | 0.94, 2.93 |
| Protease inhibitor use | 1.73 | 1.36, 2.21 | 1.07 | 0.87, 1.38 | 1.28 | 0.85, 1.92 | 0.76 | 0.48, 1.21 |
| Abacavir use | 2.06 | 1.59, 2.67 | 1.27 | 0.95, 1.68 | 1.16 | 0.71, 1.91 | 0.83 | 0.48, 1.42 |

**Model is also adjusted for CD4 count (time-varying), BMI, CD4 count at ART initiation, VL at ART initiation, and cohort.**
Abbreviations: HR, Hazard Ratio; CI, Confidence Interval

**Supplementary Table 8.** Sensitivity analysis: demographic characteristics of participants, by ever anxiety, ever depression, or neither anxiety nor depression diagnosis.

|  |  | Ever depression | Ever anxiety | Neither anxiety nor depression |
| --- | --- | --- | --- | --- |
| Characteristic |  | N = 13,438<sup>1</sup> | N = 8,980<sup>1</sup> | N = 15,326 |
| Demographics | Age, years, median (IQR) | 43 (35, 49) | 42 (35, 49.50) | 41 (33, 49) |
|  | Male sex, n (%) | 10,799 (80.4%) | 7,505 (83.6%) | 13,042 (85.1%) |
|  | Race/ethnicity, n (%) |  |  |  |
|  | Non-Hispanic White | 6,610 (49.2%) | 4,975 (55.4%) | 5,961 (38.9%) |
|  | Non-Hispanic Black | 4,106 (30.6%) | 1,853 (20.6%) | 6,008 (39.2%) |
|  | Hispanic | 1,917 (14.3%) | 1,525 (17%) | 2,294 (15%) |
|  | Other/unknown | 805 (6%) | 627 (7%) | 1,063 (6.9%) |
|  | HIV transmission risk group, n (%) |  |  |  |
|  | Heterosexual contact | 3,068 (22.8%) | 1,713 (19.1%) | 4,187 (27.3%) |
|  | MSM | 7,636 (56.8%) | 5,671 (63.2%) | 8,660 (56.5%) |
|  | PWID | 2,083 (15.5%) | 1,161 (12.9%) | 1,450 (9.5%) |
|  | Other/unknown | 651 (4.8%) | 435 (4.8%) | 1,029 (6.7%) |

|  |  | Ever anxiety | Ever depression | Neither anxiety nor depression |
| --- | --- | --- | --- | --- |
| Characteristic |  | N = 8,980<sup>1</sup> | N = 13,438<sup>1</sup> | N = 15,326 |
| Traditional CVD risk factors | Smoking, n (%) | 5,937 (70.7%) | 8,932 (73.6%) | 7,607 (59.8%) |
|  | Unknown<sup>2</sup> | 581 | 1,302 | 2,608 |
|  | At-risk alcohol use, n (%) | 3,553 (39.6%) | 5,690 (42.3%) | 3,728 (24.3%) |
|  | Cocaine use, n (%) | 990 (11.0%) | 1,856 (13.8%) | 937 (6.1%) |
|  | Cannabis use, n (%) | 1,116 (12.4%) | 1,672 (12.4%) | 612 (4%) |
|  | Hypertension, n (%) | 3,212 (35.8%) | 4,996 (37.2%) | 4,042 (26.4%) |
|  | Elevated total cholesterol, n (%) | 2,249 (25%) | 3,297 (24.5%) | 2,708 (17.7%) |
|  | Statin use, n (%) | 2,322 (25.9%) | 3,471 (25.8%) | 2,740 (17.9%) |
|  | Elevated total cholesterol OR statin use, n (%) | 3,363 (37.4%) | 5,058 (37.6%) | 4,253 (27.8%) |
|  | Chronic kidney disease, n (%) | 965 (10.7%) | 1,653 (12.3%) | 1,416 (9.2%) |
|  | Diabetes, n (%) | 993 (11.1%) | 1,684 (12.5%) | 1,404 (9.2%) |

|  |  | Ever anxiety | Ever depression | Neither anxiety nor depression |
| --- | --- | --- | --- | --- |
| Characteristic |  | N = 8,980<sup>1</sup> | N = 13,438<sup>1</sup> | N = 15,326 |
| HIV-related risk factors | CD4 at ART initiation, median (IQR) | 320 (164.50, 510) | 294.00 (135, 480) | 301.00 (129, 502) |
|  | Unknown<sup>2</sup> | 1,994 | 3,207 | 3,863 |
|  | VL at ART initiation, median (IQR) | 23,200 (500, 108,183.00) | 27,288.50<br>(1,216, 126,486.00) | 25,335 (500, 121,000.00) |
|  | Unknown<sup>2</sup> | 2,114 | 3,472 | 4,047 |
|  | AIDS diagnosis, n (%) | 1,392 (15.5%) | 2,351 (17.5%) | 2,423 (15.8%) |
|  | HCV infection, n (%) | 1,406 (16.4%) | 2,490 (19.5%) | 2,004 (14.2%) |
|  | Unknown<sup>2</sup> | 408 | 692 | 1,169 |
|  | Protease inhibitor use, n (%) | 4,881 (54.4%) | 7,889 (58.7%) | 7,217 (47.1%) |
|  | Abacavir use, n (%) | 3,239 (36.1%) | 5,104 (38.0%) | 4,642 (30.3%) |

<sup>1</sup> **6,416 (20.4%)** have both anxiety and depression
<sup>2</sup> Unknown values were missing and imputed for the analysis
Abbreviations: IQR, interquartile range; n, number of individuals; HIV, human immunodeficiency virus; MSM, men who have sex with men; PWID, people who inject drugs; CVD, cardiovascular disease; ART, antiretroviral therapy; VL, viral load; AIDS, acquired immunodeficiency syndrome; HCV, hepatitis C virus.

## Supplementary Figure

**Supplementary Figure 1.**
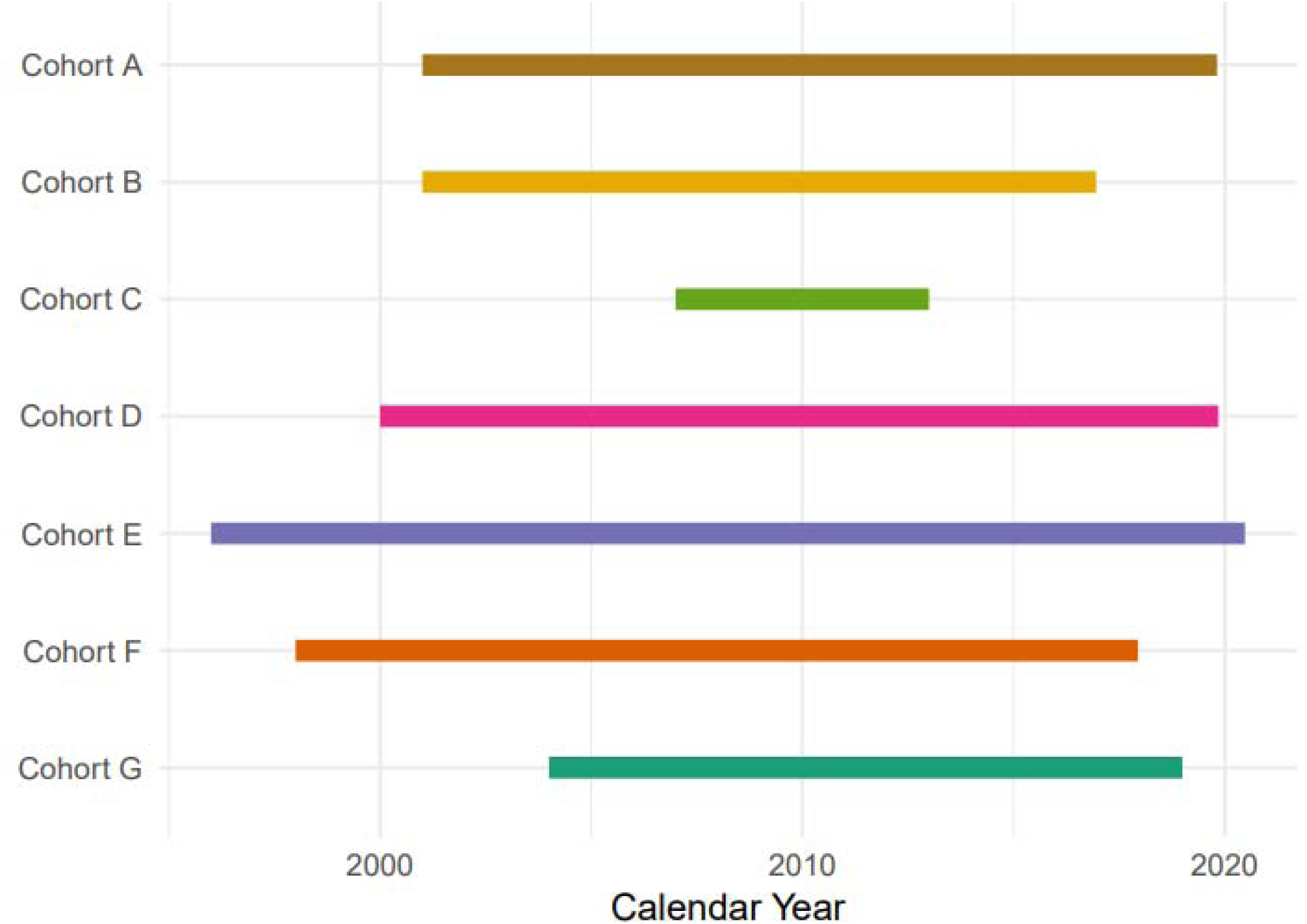
Myocardial infarction observation windows for the 7 clinical cohorts that contributed data to this nested study. Each bar shows the years of the observation window for one of the 7 contributing cohorts.

**Supplementary Figure 2.**
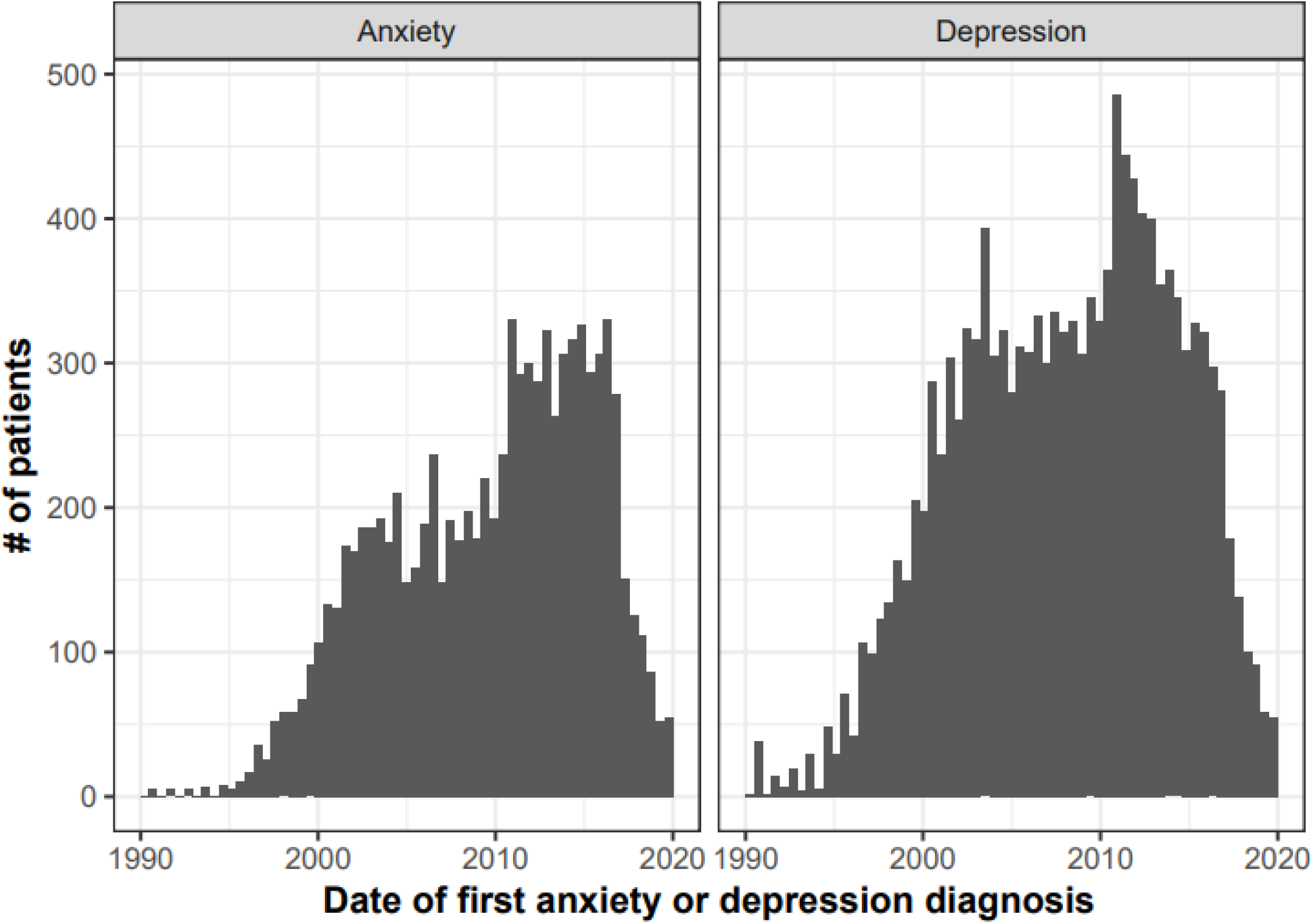
Incident diagnoses of anxiety and depression among people with HIV in NA-ACCORD from 1990 to 2020. The left panel shows the frequency distribution of incident anxiety diagnoses, while the right panel presents the frequency distribution of incident depression diagnoses. Both figures show a gradual increase in the number of diagnoses starting from the late 1990s until the mid-2000’s with a further increase in 2010, when routine screening for mental health illnesses was implemented in some clinics. After 2010, there is a decline in the number of incident diagnoses. The vertical axis represents the number of people with HIV, and the horizontal axis represents the date of the first mental health diagnosis.

**Supplementary Figure 3.**
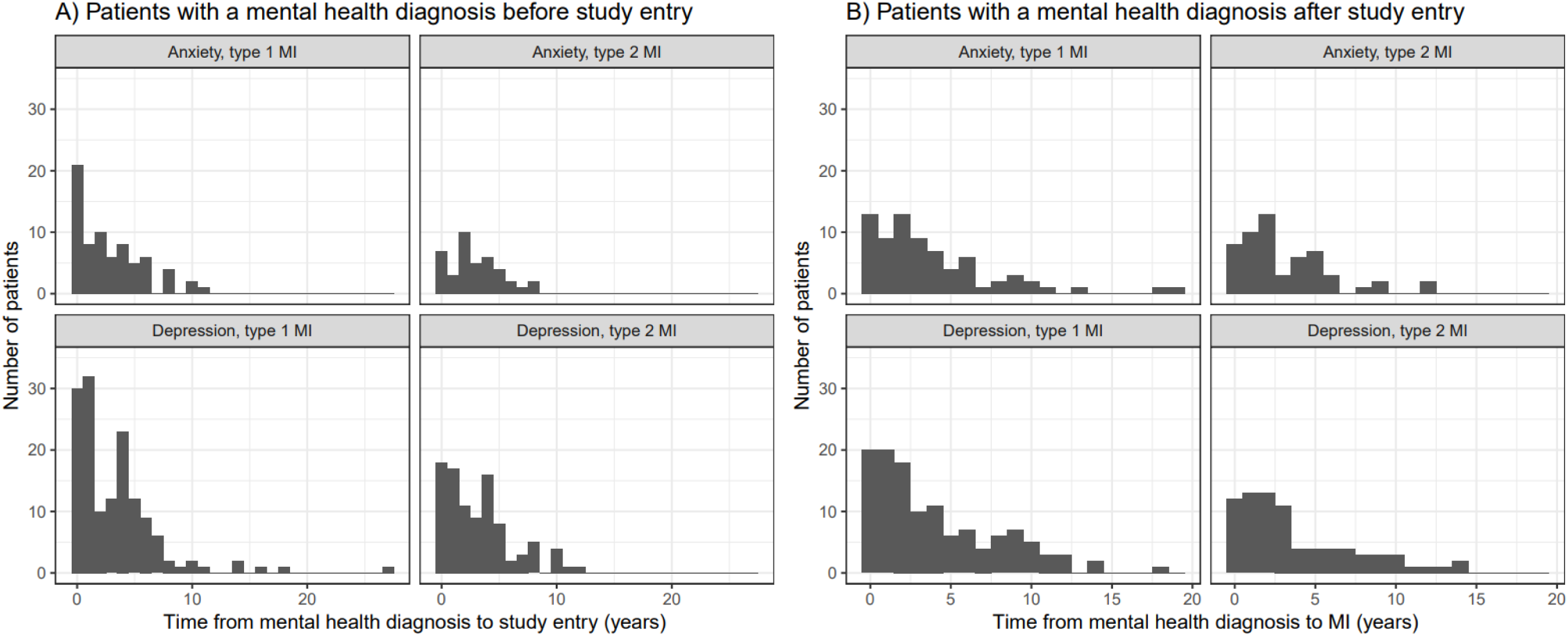
Time from diagnosis of anxiety or depression until incident Type 1 or Type 2 myocardial infarction (MI). Panel A shows the distribution of time from mental health diagnosis to study entry for participants who had a diagnosis of depression or anxiety prior to study entry. Panel B shows the distribution of time from mental health diagnosis to incident MI for participants who received a new diagnosis of depression or anxiety after study entry. Of the 272 people diagnosed with depression, 149 were diagnosed before study entry (median 2.51 years before study entry [IQR, 0.60-4.55 years]), and 123 were diagnosed on or after study entry (median 2.91 years before MI [IQR, 1.10-6.36 years]). Of the 148 people diagnosed with anxiety, 71 were diagnosed before study entry (median 2.00 years before MI [IQR, 0.43-4.46 years]), and 73 were diagnosed on or after study entry (median 2.80 years before MI [IQR, 1.11-5.47 years]). Of the 175 people diagnosed with depression, 96 were diagnosed before study entry (median 2.57 years before study entry [IQR, 0.75-4.51 years]), and 79 were diagnosed on or after study entry (median 2.63 years before MI [IQR, 1.19-5.72 years]). Of the 95 people diagnosed with anxiety, 40 were diagnosed before study entry (median 2.46 years before MI [IQR, 1.42-4.20 years]), and 55 were diagnosed on or after study entry (median 2.17 years before MI [IQR, 1.11-4.54 years]).

**Supplementary Figure 4.**
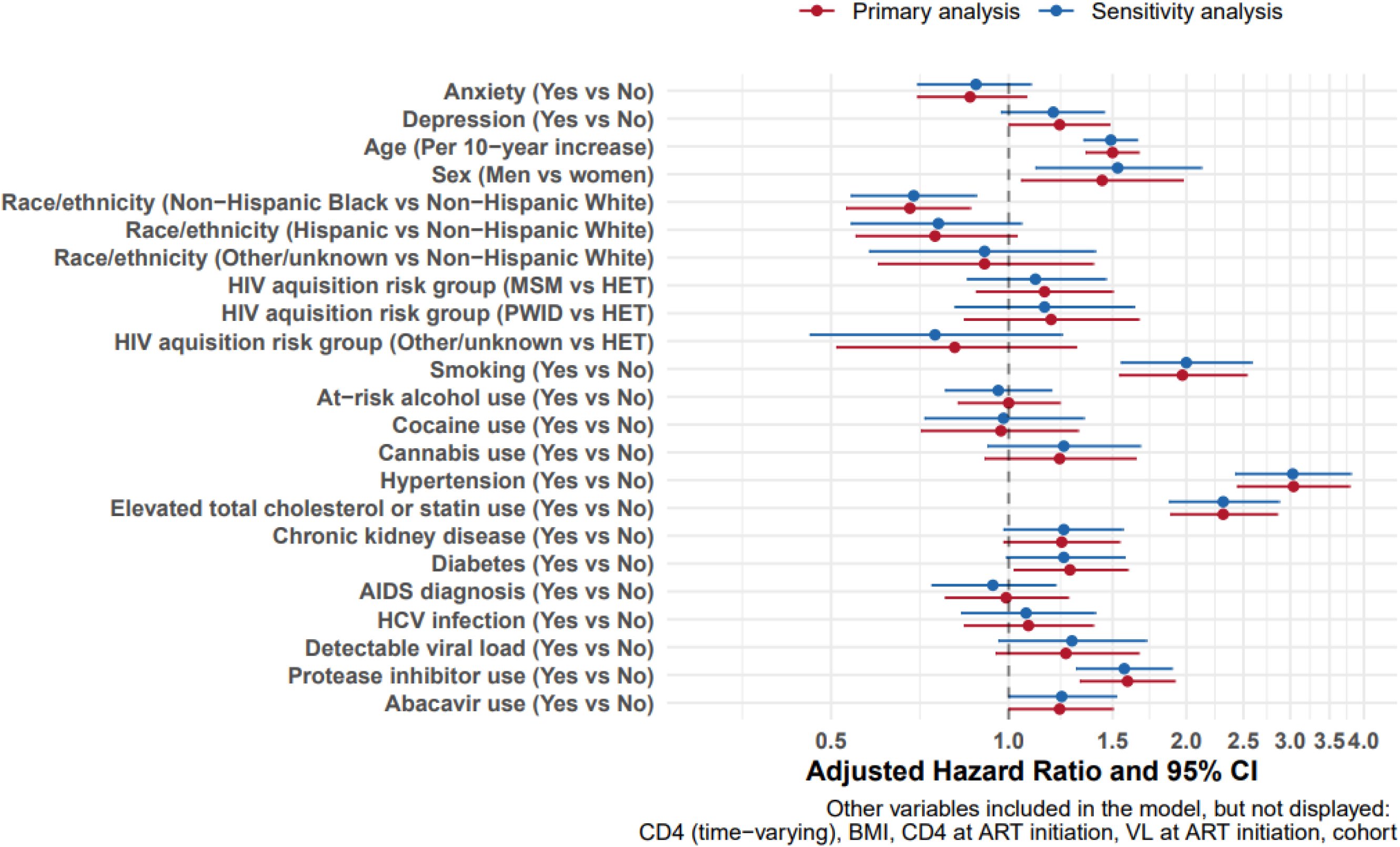
Combined primary analysis and sensitivity analysis forest plot exhibiting adjusted hazard ratios and 95% confidence intervals for associations with Type 1 myocardial infarction (MI). Forest plot presenting the adjusted hazard ratios (HR) and 95% confidence intervals (CI) for various factors associated with incident Type 1 MI, analyzed separately for the primary analysis (blue) and sensitivity analysis (red). The vertical dashed line indicates a hazard ratio of 1.0, signifying no effect. Points to the right suggest increased risk, while points to the left suggest decreased risk.

**Supplementary Figure 5.**
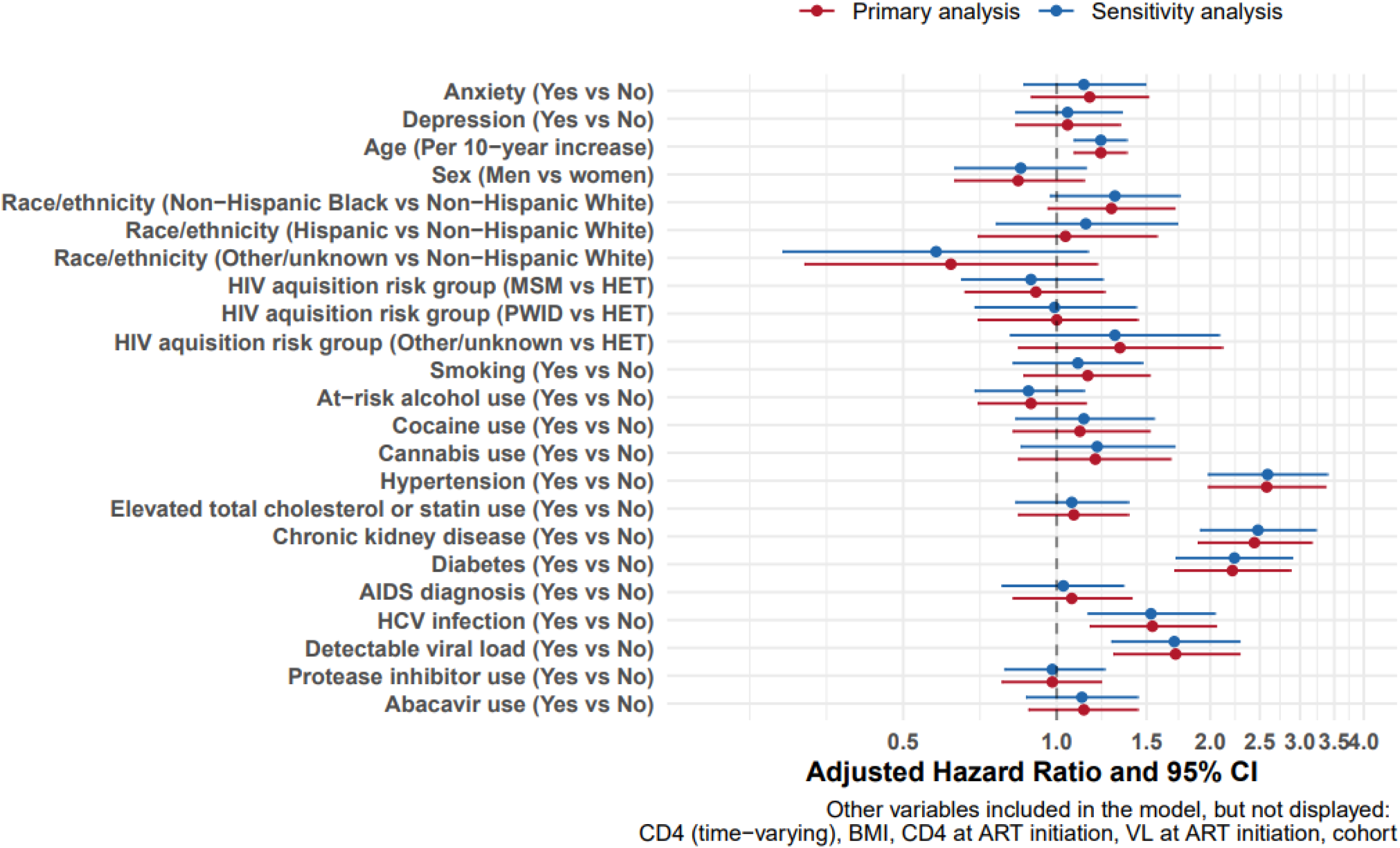
Combined primary analysis and sensitivity analysis forest plot exhibiting adjusted hazard ratios and 95% confidence intervals for associations with Type 2 myocardial infarction (MI). Forest plot illustrating the adjusted hazard ratios (HR) and 95% confidence intervals (CI) for various factors impacting incident Type 2 MI, analyzed separately for the primary analysis (blue) and sensitivity analysis (red). The vertical dashed line marks a hazard ratio of 1.0 (no effect). Points to the right of this line suggest increased risk, while points to the left suggest decreased risk.

